# MedDx, a training-free AI phenotyping model, identifies alcohol use disorder in cirrhosis with hepatologist-level agreement

**DOI:** 10.64898/2026.11.25.26368618

**Authors:** Haining Wang, Yao Chen, Maaz Arif, Deborupa Pal, Chenxi Xiong, Hongshan Liu, Suthat Liangpunsakul, Archita P. Desai, Wanzhu Tu, Samer Gawrieh, Jing Su

## Abstract

Alcohol use disorder (AUD) is often under-documented in patients with cirrhosis, which delays treatment. Expert chart review of clinical notes can identify under-documented AUD, but is not scalable. We propose MedDx, an open-weight, training-free, and engine-agnostic language model, to phenotype AUD using clinical notes in patients with cirrhosis. We benchmarked MedDx on a 485-patient study cohort sampled from the 14,406-patient Indiana Cirrhosis Registry, using a chart-review-based reference as ground truth. MedDx agreed with the ground truth at three-class Cohen’s κ 0.92, against 0.67 for the AUD diagnosis code. It agreed with one hepatologist at 0.85 on all 485 charts and with a second at 0.82 on 186 charts, on which the two hepatologists agreed at 0.87. Weighted to the registry, its sensitivity was 98% and specificity 94%, against 63% and 96% for the AUD diagnosis code, and 61% of an estimated 6,223 patients with AUD had no AUD diagnosis by the first cirrhosis diagnosis. MedDx is openly available as an off-the-shelf, equitable, and auditable tool to identify AUD.

---

Alcohol use disorder (AUD), a treatable clinical condition^1^, affects more than one in ten US adults^2^. In patients with cirrhosis, timely AUD treatment is associated with fewer hepatic decompensations and lower mortality^3,4^. Diagnosing AUD requires clinicians to interview patients against the DSM-5-TR criteria^5^ and document positive cases using International Classification of Diseases (ICD) diagnosis codes^6^ for AUD in electronic health records (EHRs). However, although AUD-related evidence is often documented in clinical notes, a formal diagnosis is often missing^7,8^. Under-documentation of AUD has become a major barrier to timely AUD interventions and efficient management of alcohol-associated liver diseases (ALD)^9^, especially in advanced cases such as cirrhosis. Furthermore, completeness of AUD diagnosis is associated with demographic characteristics, socioeconomic conditions, and hospital type. Prior studies report less complete AUD diagnosis in women, Black patients, disadvantaged groups, and patients served by safety-net hospitals, contributing to the health disparity of cirrhosis management^10–12^. Expert chart reviews of EHR, especially clinical notes, can identify under-documented AUD. However, this approach is time-consuming, inconsistent due to clinicians’ experience, and not scalable.

The rapid development of artificial intelligence (AI) models, especially general large language models (LLMs), provides a promising solution for scalable and consistent chart review^13–15^ of social and behavioral information in clinical notes^16,17^ and phenotyping substance abuse^18–21^. Recently, a fine-tuned transformer model^8^ digests discharge summaries to identify AUD cases. Also, a rule-based natural language processing model retrieves key evidence from notes using keyword search to identify patients with AUD^22^. A trustworthy and clinically implementable LLM measurement instrument^23–25^ should align with expert-generated ground truth, demonstrate consistent performance across patient groups with different demographic features and socioeconomic conditions and across hospital types, distinguish uncertain cases from negative and positive cases, be auditable and traceable for evidence supporting each classification, and be free of hallucination. However, to our knowledge, no LLM-based solution has been developed and benchmarked as an instrument for identifying AUD using clinical notes in patients with cirrhosis.

In this work, we developed MedDx, an open-weight, off-the-shelf AI instrument for clinical notes-based AUD phenotyping among patients with cirrhosis. The model performance was benchmarked on the Indiana Cirrhosis Registry (ICR)^26^, a statewide registry of 14,406 patients with cirrhosis built on the Indiana Network for Patient Care (INPC)^27^, using a 485-patient study cohort^28^ with ground truth generated through hepatologist-adjudicated chart review. MedDx demonstrates comparable performance to clinician chart reviews, and demonstrates consistent accuracy across patient groups and hospital systems, whereas the diagnosis’s accuracy varies widely. As an agnostic AI solution, MedDx supports mainstream open-weight LLM engines, and with ten of the 11 engines tested it agreed with the ground truth more closely than the AUD diagnosis did. As a trustworthy AI model, MedDx is auditable, traceable, and honest about uncertain cases, and 98% of its supporting quotations are verbatim in the chart. We estimate that 43% of patients in the Indiana Cirrhosis Registry (6,223 of 14,406) had AUD, of whom 61% carried no AUD diagnosis at the first cirrhosis diagnosis and two-fifths carried no alcohol-related diagnosis of any kind. MedDx’s classifications, weighted the same way, give 6,466 patients with AUD.

## Results

MedDx and the AUD diagnosis are compared throughout, each against the ground truth provided by hepatologists who read the chart (Fig. 1a). MedDx and the hepatologists read each patient’s clinical notes, those retrieved within 18 months either side of the patient’s study-cohort index date (Methods), and answer on the same six-grade scale, which the analysis collapses into three classes: AUD (AUD+), no AUD (AUD−), and indeterminate (Supplementary Table 1). A patient is indeterminate when the answer is uncertain or the notes do not contain enough data. The AUD diagnosis is read from the structured record and not from the notes. A patient with an ICD code for AUD (F10.x or an ICD-9 equivalent) recorded at any time in the structured record, which runs from 1 January 2015 to 10 July 2023, is AUD+ and a patient without one is AUD−, so the diagnosis is never indeterminate.

**Fig. 1.**
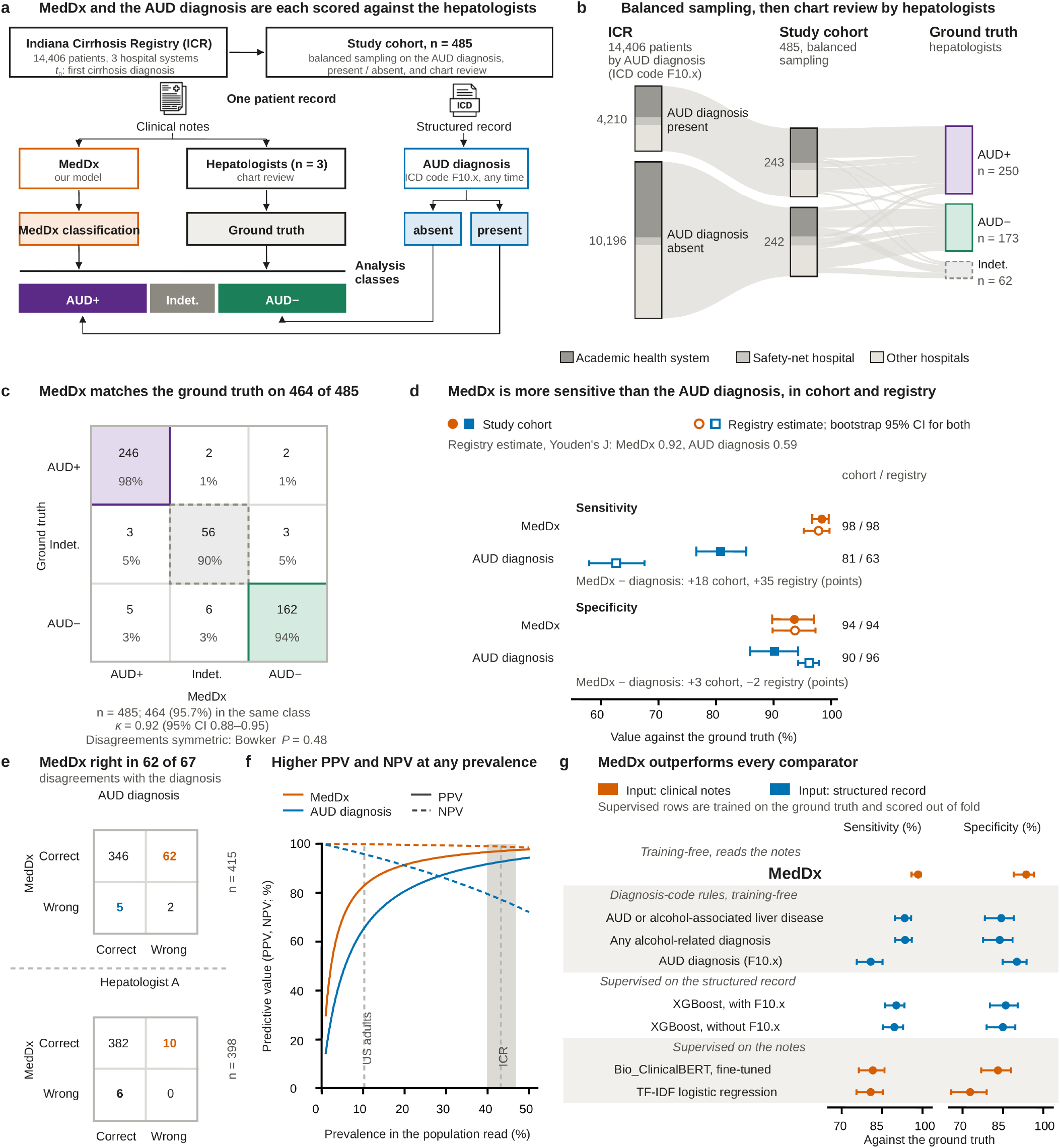
MedDx agrees closely with hepatologist chart review and identifies the patients with AUD whom the AUD diagnosis misses. Blue, the AUD diagnosis; orange, MedDx; ink, the hepatologists. Plum, AUD+; green, AUD−; gray, indeterminate. Sensitivity and specificity are computed over all 423 charts with an AUD+ or AUD− ground truth; a chart that MedDx left indeterminate stays in the denominator. **a**, Study design. MedDx (Fig. 4a) and the hepatologists, whose classification is the ground truth, read the same clinical notes (Extended Data Figs. 1 and 2); the AUD diagnosis is an ICD code recorded at any time in the structured record and is never indeterminate. **b**, Within each column, node heights are proportional to patients, and the bands are the reviewed charts; the segments of the first two columns are the three hospital systems, and each band leaves the segment of its own system. **c**, Ground truth (rows) against MedDx (columns), count and row percentage per cell; κ is computed on the 428 charts on which neither returned not enough data. Six grades: Supplementary Table 1. **d**, Sensitivity and specificity of MedDx and of the AUD diagnosis in the study cohort and in the registry estimate (Methods). **e**, Charts by whether MedDx (rows) and a comparator (columns), the AUD diagnosis or hepatologist A, matched the ground truth. The discordant counts carry the color of the classifier that was right, in bold, and the test uses only those (Methods). The 415 and 398 denominators exclude charts on which MedDx or hepatologist A abstained. **f**, Positive (solid) and negative (dashed) predictive value against prevalence, from the registry-weighted rates of positive and negative classification, held fixed across prevalence. The rates of the AUD diagnosis reflect coding practice in this registry and may differ elsewhere. Broken lines mark the registry estimate of AUD prevalence in the ICR, 43%, its 95% CI shaded, and past-year AUD among US adults, 10%, shown as a low-prevalence reference. **g,** MedDx has the highest sensitivity and the highest specificity of the eight classifiers scored on the same 423 charts. The classifiers are in four groups, ordered by Youden’s J within each: sensitivity (left) and specificity (right) with Wilson 95% CI. Of the 48 ground-truth-positive patients who carry no AUD diagnosis, MedDx identified 46 (95.8%), the strongest comparator on the structured record 68.8%, and the two supervised text models 64.6% and 68.8%. Specifications: Methods and Supplementary Tables 15, 18 and 19.

The study cohort is 485 patients drawn from the ICR in balanced numbers with and without an AUD diagnosis (Fig. 1b; Extended Data Table 1). Hepatologist A reviewed every chart, and hepatologist B independently reviewed a random subsample of 186 charts. A chart underwent a second reading by hepatologist C when MedDx, run with any of three LLM engines, or hepatologist B placed the patient in a different class than hepatologist A. When hepatologists A and C disagreed, hepatologist D decided (Methods; Extended Data Figs. 1 and 2). No hepatologist saw any MedDx output at any stage. Hepatologist B’s classifications are not part of the ground truth. The ground truth is AUD+ on 250 charts, AUD− on 173, and indeterminate on 62. The 423 charts with an AUD+ or AUD− ground truth are the denominator for sensitivity, specificity and predictive value.

**Fig. 2.**
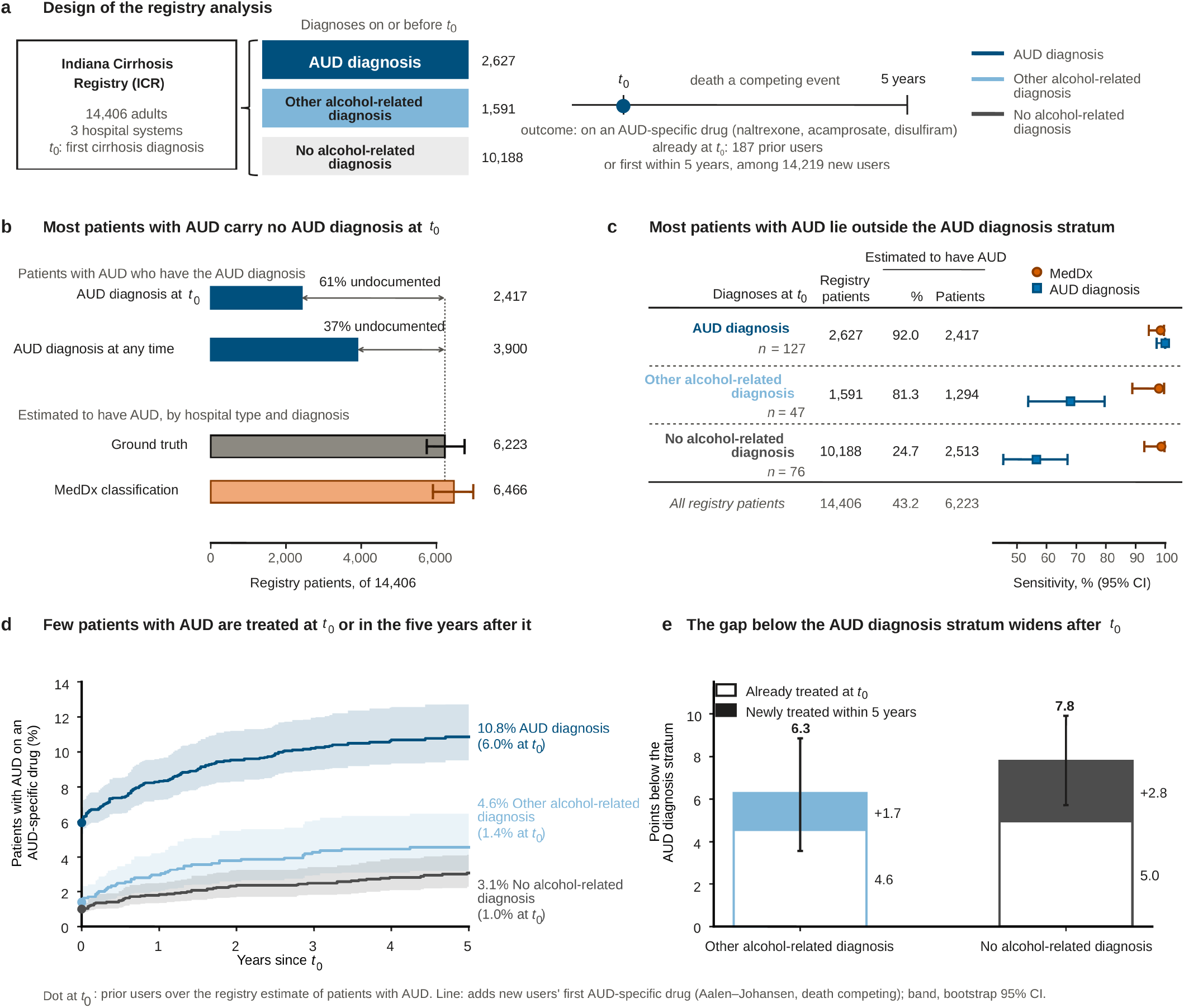
Of the registry patients estimated to have AUD, 61% carry no AUD diagnosis at the first cirrhosis diagnosis, and those with no alcohol-related diagnosis are the least often treated. *t*_0_ is the first cirrhosis diagnosis, and diagnoses on or before *t*_0_ define the three strata. The registry estimate weights each reviewed chart with an AUD+ or AUD− ground truth to the registry patients it represents; intervals are 95% percentile bootstrap intervals over the reviewed charts (Methods). **a,** Design. Patients with an AUD-specific drug recorded before *t*_0_ (187: 144, 18 and 25 in the three strata) are the prior users, counted as treated at *t*_0_ in d and e; the 14,219 new users are followed from *t*_0_ for a first naltrexone, acamprosate or disulfiram, with death as a competing event. **b,** Registry patients estimated to have AUD who have the AUD diagnosis at *t*_0_ and at any time in the structured record, against all patients estimated to have AUD from the ground truth (dotted line) and from MedDx’s classifications of the same charts with the same weights. Undocumented is the share of patients estimated to have AUD who do not have the diagnosis. **c,** The three strata at *t*_0_. At the right, sensitivity of MedDx and of the AUD diagnosis (counted at any time) on the reviewed charts of each stratum; *n* is the ground-truth-positive charts. The sample over-represents patients diagnosed after *t*_0_, so these sensitivities are not registry values. Sensitivity by hospital system: Supplementary Table 7. **d,** The share of patients with AUD in each stratum on an AUD-specific drug. The dot at *t*_0_ is the prior users over the registry estimate of patients with AUD; the line adds the cumulative incidence of a first drug after *t*_0_ among every new user of the stratum, each recipient counted as a patient with AUD (Aalen–Johansen, death competing). The curves for every new user, with numbers at risk, are in Extended Data Fig. 7. **e,** Percentage points by which each stratum lies below the AUD diagnosis stratum in d: among prior users at *t*_0_ (open) and added over the five years after it (filled; 1.7 points, 95% CI −0.2 to 3.5, and 2.8 points, 1.3–4.2). The bar is the 95% CI of the total, 3.6–8.9 and 5.7–9.9 points. The shares at one year and the ratios are in Extended Data Table 2.

### Agreement with expert chart review

MedDx placed 464 of 485 patients in the same class as the ground truth, and the three-class Cohen’s κ was 0.92 (95% confidence interval (CI) 0.88–0.95) (Fig. 1c). It classified two patients with an AUD+ ground truth as AUD− and five with an AUD− ground truth as AUD+. In the remaining 14, one classification was indeterminate and the other was not: MedDx’s in 8 and the ground truth’s in 6 (six grades: Supplementary Tables 1 and 2). The disagreements favored neither class (Bowker’s test of symmetry, *P* = 0.48). Against the hepatologists’ first-pass readings, before adjudication, MedDx reached κ 0.85 (95% CI 0.81–0.90) with hepatologist A on every chart and 0.82 (0.73–0.90) with hepatologist B on the 186 charts hepatologist B read. On those 186 charts, hepatologists A and B reached 0.87 (0.79–0.94) with each other (Supplementary Table 3; Supplementary Note 1).

The study cohort is balanced on the AUD diagnosis, whereas the registry includes 4,210 patients with it and 10,196 without, so we weighted each reviewed chart to the registry patients it represents (Fig. 1d; Methods). Weighted to the registry, MedDx has a sensitivity of 98% (95% CI 95–100) and a specificity of 94%, each within one percentage point of its value on the reviewed charts, whereas the sensitivity of the AUD diagnosis falls from 81% on the reviewed charts to 63% (58–68), with a specificity of 96% (Supplementary Table 4; Supplementary Note 1). Youden’s J^29^, sensitivity plus specificity minus one, weighted to the registry is 0.92 for MedDx and 0.59 for the AUD diagnosis. Against the verification-independent ground truth, built from the 186 charts that two hepatologists read independently so that no chart was selected by MedDx, registry-weighted specificity was 88% for MedDx and 97% for the AUD diagnosis (Methods).

When MedDx and the AUD diagnosis disagreed, MedDx was usually correct. Of the 67 charts in which one was right and the other wrong, MedDx was correct in 62 (McNemar *P = 7.8 × 10−12, χ2 = 4C.8*; Fig. 1e). Weighted to the registry, MedDx’s positive predictive value was 97% (95% CI 94–99) and its negative predictive value 99% (97–100), compared with 93% (89–96) and 77% (72–82) for the AUD diagnosis (Fig. 1f).

Among three diagnosis-code rules, a gradient-boosted model on the structured record and two supervised text models, all scored against the same ground truth, none recovered as many of the patients the AUD diagnosis misses as MedDx did (Fig. 1g). Of the 48 ground-truth-positive patients who carry no AUD diagnosis, MedDx identified 46 (95.8%), whereas the strongest comparator on the structured record and a fine-tuned Bio_ClinicalBERT^30^ each recovered 68.8%. The code rules and the fine-tuned BERT correspond to the two approaches earlier applied to AUD^8,22^.

### Under-documentation and under-treatment of AUD

An estimated 43.2% of registry patients (95% CI 39.9–46.8), 6,223 of 14,406, have AUD (Fig. 2a,b). The registry estimate weights each reviewed chart with an AUD+ or AUD− ground truth to the registry patients it represents (Methods; Extended Data Figs. 3 and 4). Projecting MedDx’s classifications of the same charts with the same weights yields 6,466 (5,906–6,981).

Of the 6,223 registry patients estimated to have AUD, 61% have no AUD diagnosis at the time of the first cirrhosis diagnosis, and 37% have no AUD diagnosis at any time in the structured record (Fig. 2b). Diagnoses on or before the cirrhosis diagnosis define three strata: the AUD diagnosis stratum (2,627 patients), the other alcohol-related diagnosis stratum (1,591), defined mainly by codes for alcohol-associated liver disease (Supplementary Note 2), and the no alcohol-related diagnosis stratum (10,188). An estimated 2,417, 1,294 and 2,513 patients with AUD are in the three strata (Extended Data Table 2), so two-fifths of patients with AUD carry no alcohol-related diagnosis of any kind at the cirrhosis diagnosis. On the reviewed charts, MedDx identified 98%, 98% and 99% of the patients with AUD in the three strata (Fig. 2c).

Patients with AUD who had no AUD diagnosis at the time of cirrhosis diagnosis were treated less often. By five years, 10.8% (95% CI 9.4–12.7) of patients with AUD in the AUD diagnosis stratum were on naltrexone, acamprosate, or disulfiram, compared with 4.6% (3.4–6.6) in the other alcohol-related diagnosis stratum and 3.1% (2.2–4.2) in the no alcohol-related diagnosis stratum (Fig. 2d). Most of the difference was present at the time of cirrhosis diagnosis, when 6.0%, 1.4%, and 1.0% had a drug recorded, and it widened afterwards (Fig. 2e; Extended Data Table 2).

### Accuracy across patient groups and hospital systems

MedDx’s sensitivity stayed within 5 percentage points of the other groups for the same variable across all 18 patient groups, with the largest difference at 2.3 points. The groups were formed by age, sex, race, urban–rural residence, neighborhood deprivation and hospital system (Methods). The sensitivity of the AUD diagnosis differed by more than 5 points in 8 of the 18 groups, and its specificity did so in 9. The largest differences were −21.0 points in sensitivity at the safety-net hospital and −20.3 points in specificity in the most deprived neighborhood quintile. MedDx’s specificity differed by more than 5 points in 4 groups, with the largest difference at 7.3 points (Fig. 3a,b; Supplementary Table 6). Three of the four intervals include zero, and the fourth is the under-50 group, where MedDx’s specificity is higher than in the rest. Plotted as operating points, the 18 MedDx groups span sensitivities of 97.1 to 100.0% and false positive rates of 0.0 to 12.5%, against 62.5 to 91.0% and 2.4 to 25.6% for the AUD diagnosis (Fig. 3d).

**Fig. 3.**
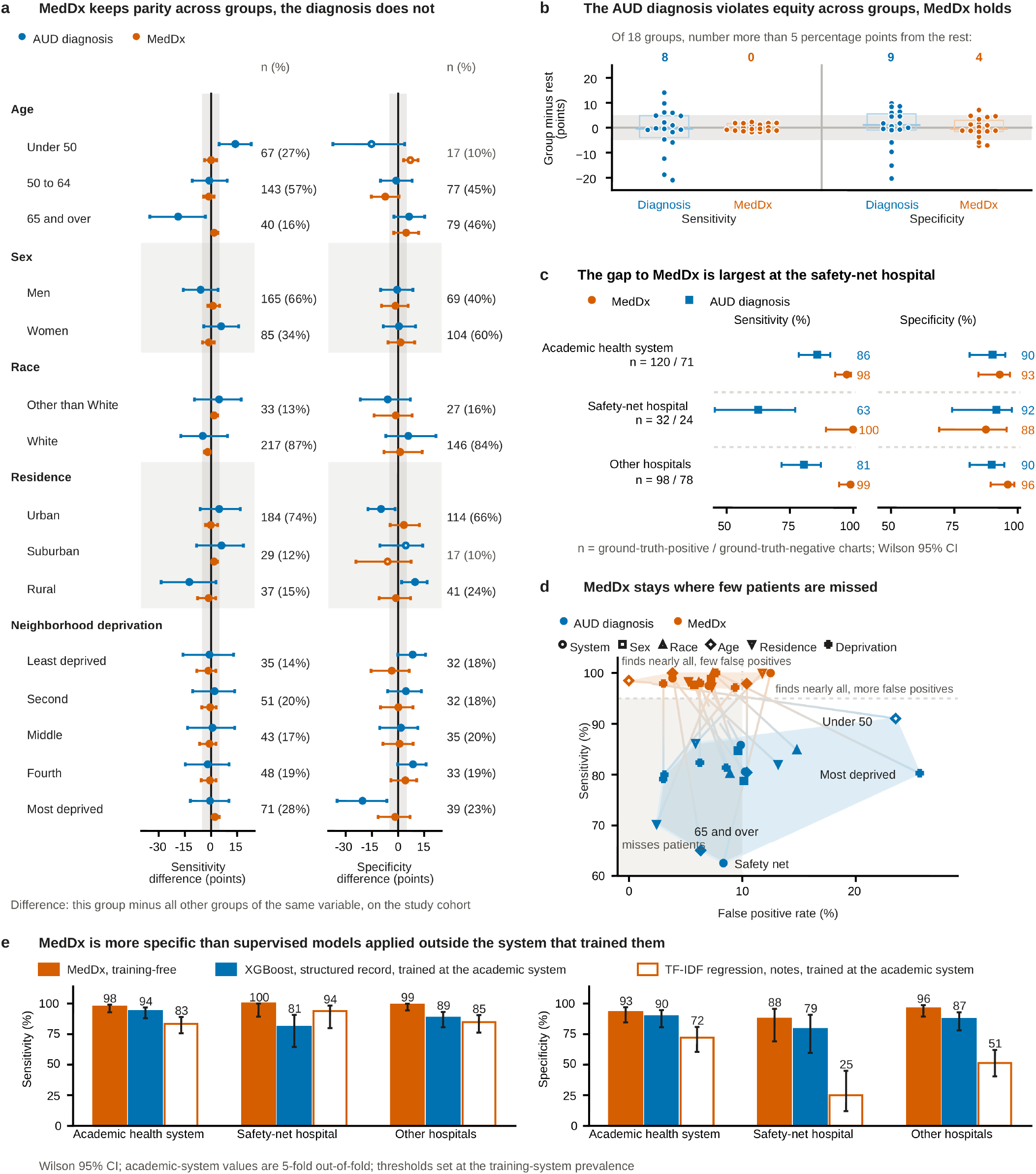
The accuracy of MedDx varies little across patient groups and hospital systems, whereas that of the AUD diagnosis varies widely. All panels use the 485 charts of the study cohort, of which 250 are ground-truth-positive and 173 ground-truth-negative. An indeterminate output stays in the denominator. Every comparison is one group versus the remaining groups of the same variable; gray shading marks a 5-point reference band. Intervals are percentile bootstrap 95% intervals, resampled within sampling stratum. **a**, Sensitivity and specificity of each group minus the rest, in percentage points, for the AUD diagnosis (blue) and MedDx (orange); zero indicates no difference. Beside each half, the charts in the group show n (%) of the ground-truth-positive (left) or ground-truth-negative (right) charts; an open marker indicates fewer than 20. **b**, All 18 group differences for a and c for the AUD diagnosis and for MedDx, one point per group, for sensitivity (left) and specificity (right). The box is the interquartile range, the line across it the median; the band is 5 points, and the number above each column counts the groups outside it. The registry-scale sensitivity of the diagnosis by group is in Supplementary Fig. 2; the same spread for every engine and skill variation is in Supplementary Fig. 3. **c**, Sensitivity and specificity by hospital system, as absolute values with Wilson 95% CI; n is the ground-truth-positive and ground-truth-negative charts in each system. **d**, The same groups as operating points, with hospital system added, are colored as in a and shaped by variable, with the convex hull of the points of each; a line shaded from blue to orange joins the diagnosis’s point to MedDx’s for the same group. The hulls cover 399 square percentage points for the diagnosis and 23 for MedDx. **e,** MedDx is more specific than two supervised models trained at the academic health system (191 AUD+ or AUD− charts) and applied unchanged to the safety-net hospital and the other hospitals: the gradient-boosted model over the structured record with the AUD diagnosis among its inputs, and the TF-IDF logistic regression over the notes. Bars are sensitivity (left) and specificity (right) with Wilson 95% CI; the academic-system values are 5-fold out-of-fold on the training charts. Thresholds were set at the academic health system and carried unchanged (Methods). Values: Supplementary Table 8.

The gain from reading the notes was greatest at the safety-net hospital. The paired gain ranged from +11.7 percentage points at the academic health system to +37.5 at the safety-net hospital. The sensitivity of the AUD diagnosis was 85.8% at the academic health system, 80.6% at the other hospitals and 62.5% at the safety-net hospital. MedDx’s sensitivity was 97.5%, 99.0% and 100% in the same three. Its agreement with the ground truth was similar across all three, with κ 0.89 (95% CI 0.83–0.95), 0.96 (0.91–0.99), and 0.89 (0.77–1.00) (Fig. 3c; Supplementary Table 7). At the safety-net hospital, MedDx, which is training-free, held a sensitivity of 100% and a specificity of 88%, a higher specificity than either of two supervised models trained at the academic health system and applied there unchanged, 79% and 25% (Fig. 3e; Supplementary Table 8). Agreement was also similar across the three calendar periods of the cirrhosis diagnosis (Supplementary Fig. 1).

MedDx and the hepatologists were indeterminate on the same 56 charts. The rate of indeterminate classification varied by group in the same way for both, rising from urban to rural residence (Supplementary Table 9). At registry scale, the sensitivity of the AUD diagnosis varies by age and recorded race, highest under 50 and among patients recorded as other than White. Most of the patients it misses are in the largest groups, those recorded as White and those aged 50 to 64 (Supplementary Fig. 2; Supplementary Table 10). On the reviewed charts, the spread of sensitivity across groups stays narrow for eight of the 11 engines and for MedDx under each skill variation (Supplementary Fig. 3; Supplementary Table 11).

### Robustness and auditability

MedDx has three components, a skill, a harness and an open-weight language model engine (Fig. 4a; Methods). The skill is the classification instructions, which mirror how the hepatologists decide. It has four rules, each sufficient for AUD+, and requires every classification to name its rule findings, the note type or section it drew on, and a verbatim quotation. The harness is the fixed procedure that runs the skill on the engine, one patient at a time. The engine is any open-weight language model, and Qwen3.6-27B is the primary engine. We varied the engine, the decoding temperature and the skill’s wording: ten other engines, two temperatures and two paraphrases.

**Fig. 4.**
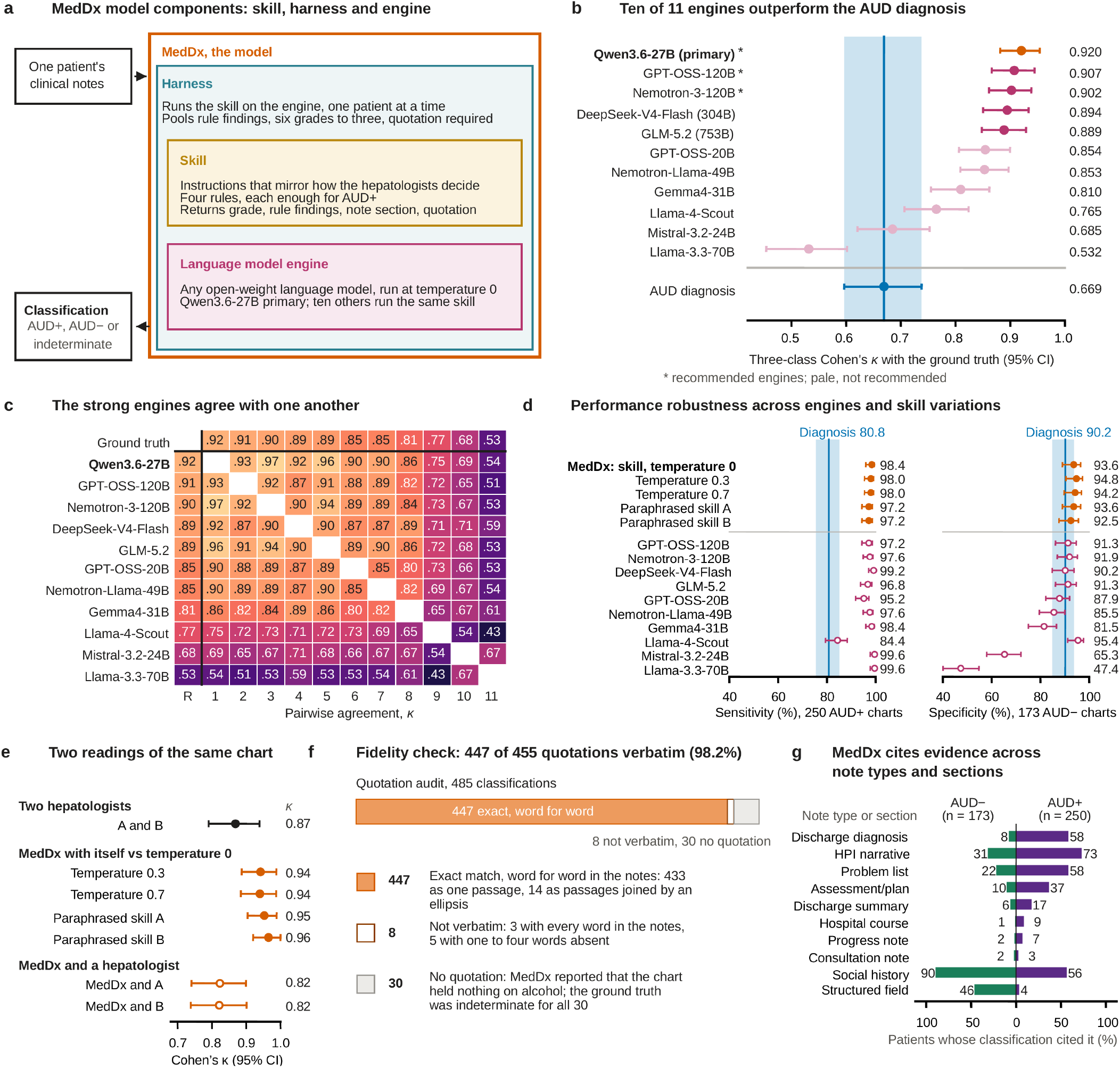
MedDx is robust across LLM engines and skill variations, and its supporting quotations are faithful to the chart. Agreement is three-class Cohen’s κ, 2,000-sample percentile bootstrap intervals over patients; a chart is dropped when either classifier returned not enough data (Methods). The primary engine, Qwen3.6-27B, is bold. **a**, MedDx has three components, the skill, the harness and the engine (Methods). **b**, Agreement with the ground truth for the 11 engines on the 485 charts and, below the rule, for the AUD diagnosis, whose κ runs through the engine rows as a line and band. Asterisks mark the three recommended engines, pale markers the engines not recommended. **c**, Pairwise agreement among 11 LLM engines, in the order of b; R is the ground truth. **d**, Sensitivity (left, 250 ground-truth-positive charts) and specificity (right, 173 ground-truth-negative charts), Wilson 95% CI, for MedDx under the skill and its four variations (filled) and the other ten engines (open); the blue line and band are the AUD diagnosis. **e**, Agreement between two readings of the same chart on the 186 charts hepatologist B read: hepatologists A and B, MedDx against each hepatologist, and MedDx against itself under two temperatures and two paraphrases of the skill. **f**, Audit of the quotations of the 485 classifications against the notes (Methods). **g,** The ten note types or sections MedDx cited most often: the share of ground-truth AUD− (green, *n* = 173) and AUD+ (plum, *n* = 250) patients whose classification cited each. The structured field is a form field inside the note, such as the social history box.

With ten of the 11 engines, MedDx agreed more closely with the ground truth than the AUD diagnosis did (Fig. 4b; Supplementary Table 12). The engines came from eight developers and read the same 485 charts under the same skill and harness. For eight engines, the lower bound of the 95% confidence interval exceeded the upper bound for the AUD diagnosis. Agreement ranged from κ 0.53 (95% CI 0.45–0.60) for Llama-3.3-70B to 0.92 (0.88–0.95) for Qwen3.6-27B, compared with 0.67 (0.60–0.74) for the AUD diagnosis. We recommend the three engines with the highest agreement, Qwen3.6-27B, GPT-OSS-120B and Nemotron-3-120B (rank under resampling, Supplementary Table 13).

The leading engines also agreed closely with one another, and the lowest pairwise κ values all involved at least one of the three lowest-ranked engines (Fig. 4c). Across all 11 engines, pairwise κ ranged from 0.43 to 0.97, with a median of 0.80. On the 250 ground-truth-positive charts, ten of the 11 engines reached 95.2% to 99.6% sensitivity, compared with 80.8% for the AUD diagnosis (Fig. 4d). On the 173 ground-truth-negative charts, specificity ranged from 47.4% to 95.4%, against 90.2% for the AUD diagnosis, with the lowest values for the three engines that rarely returned AUD−.

MedDx maintained its agreement with the ground truth across the skill and its four variations, with κ ranging from 0.90 to 0.93 (Extended Data Fig. 5). The variations were the same skill at temperatures 0.3 and 0.7, and paraphrased skill A and paraphrased skill B at temperature 0. Paraphrased skill B produced the largest paired change from the skill, −0.018 (95% paired bootstrap interval −0.042 to +0.004). All four paired intervals included zero, and each skill variation changed the three-class placement of 10 to 13 of the 485 charts. On the 186 charts hepatologist B also read, MedDx kept the same class under each variation on 96.8 to 97.8% of charts (κ 0.94 to 0.96; Fig. 4e). On those charts, hepatologists A and B agreed on 91.9% (κ 0.87), and MedDx agreed with either hepatologist on 89.2 to 89.8% (κ 0.82).

MedDx’s supporting quotations can be checked against the chart: 447 of the 455 are verbatim in the source note (98.2%; 95% CI 96.6–99.2) (Fig. 4f). Of the 485 quotation fields, 433 match one passage word for word and 14 join non-adjacent passages with an ellipsis. The other eight differ from the note in formatting, and manual review found no hallucinated content in any of them. The remaining 30 fields carry no quotation because MedDx reported that the chart held nothing about alcohol, and the ground truth was indeterminate for all 30.

MedDx identified at least one rule finding in 230 of the 250 ground-truth-positive patients, most often an explicit diagnosis of AUD in the notes (201 patients). Among those 250 patients, it most often cited the history of present illness (73%), the discharge diagnosis (58%) and the problem list (58%). Among the 173 ground-truth-negative patients, it most often cited the social history (90%) and the structured field (46%) (Fig. 4g; Extended Data Fig. 6).

## Discussion

Our results show that MedDx, a language model reading clinical notes, identifies AUD in patients with cirrhosis in close agreement with hepatologists and more accurately than the ICD diagnosis code for AUD. We tested it against hepatologists’ review of 485 charts from a statewide registry of 14,406 patients with cirrhosis. Its agreement with the hepatologists was similar across the hospital systems studied, and its sensitivity varied little across patient groups. Weighting the hepatologists’ classifications to the registry, we estimate that most of its patients with AUD carried no diagnosis code for AUD at the first cirrhosis diagnosis. These results add the clinical notes as a source that work now relying on diagnosis codes can use.

MedDx kept its accuracy across hospital systems, which we attribute to two properties. First, it reads what clinicians wrote. A diagnosis code answers to billing and to local documentation practice^12,31^, whereas notes are written by clinicians for clinicians, and the gain in sensitivity from reading them was greatest at the safety-net hospital, where the diagnosis code’s sensitivity was lowest. Second, it is training-free. Phenotypes and models fitted at one institution often lose accuracy at another^32–34^, and both supervised models, trained at the academic health system, the best-resourced of the three, lost specificity when applied unchanged elsewhere. MedDx has no training step, so there is nothing to refit when it moves to another hospital.

Another registry can run MedDx as it stands and check what it returns. The skill states the hepatologists’ rule in plain language and is released in full (Supplementary Note 3; Code availability), every engine we tested has open weights, and the leading engines agreed closely with one another, which gives registries a choice among tested engines. Each classification names its rule findings, the note type or section it drew on and a verbatim quotation, so a clinician can check it against the chart, which neither a diagnosis code nor a model score allows. A fabricated quotation, a known failure of language models^35,36^, would show as text absent from the note. Where the notes held nothing about alcohol, MedDx returned indeterminate, as the hepatologists did, and the indeterminate class shows a user where the notes leave MedDx little to read^37^.

Our findings extend earlier work showing that diagnosis codes miss AUD confirmed by chart review and that clinical notes can identify the patients the codes miss^7,8,22^. In cirrhosis, earlier studies compared alcohol-related codes with the chart for alcohol-associated liver disease^38^ and found unhealthy alcohol use on screening^39^. Hepatologist adjudication in a population-based cirrhosis registry allowed us to estimate whom a cohort defined by these codes would include. At the first cirrhosis diagnosis, requiring a diagnosis code for AUD would exclude three-fifths of registry patients with AUD, and accepting any alcohol-related diagnosis code would still exclude two-fifths.

Among registry patients with AUD, uptake of AUD-specific medication was lowest in those with no alcohol-related diagnosis code at the first cirrhosis diagnosis. Low uptake is established nationally^40^ and among patients with cirrhosis and a diagnosis code for AUD^3^. Within one year of an alcohol-associated cirrhosis diagnosis, 0.8% of commercially insured patients and 9.7% of veterans with high-risk drinking started an AUD medication^4,41^, and patients with AUD and cirrhosis had lower odds of receiving one than patients with AUD and no liver disease, an adjusted odds ratio of 0.22^42^. The estimates in cirrhosis start from a recorded alcohol-related diagnosis code. Our results extend them to patients whom a cohort defined by alcohol-related diagnosis codes would miss at the first cirrhosis diagnosis.

This study has limitations. First, disagreement between MedDx and hepatologist A was one reason a chart received a second reading, so agreement with the ground truth may be optimistic. Second, the registry is built on a single statewide health information exchange and all its patients have cirrhosis, so MedDx remains to be validated in other registries and conditions. Third, some patient groups were small, and comparisons between groups should be repeated in larger samples^43,44^.

Our results underline that the AUD the structured record under-documents in cirrhosis is largely written in the notes, where MedDx reads it with similar accuracy across hospital systems. A prospective study could test whether showing clinicians the chart evidence that MedDx identifies changes who receives treatment.

## Methods

### Study design, registry and study cohort

This was a retrospective diagnostic accuracy study using clinical notes from the statewide health information exchange on which the registry is built (the Indiana Network for Patient Care^27^). Structured data came from the registry’s structured record, and the 14,406 patients with a K70, K74 or ICD-9 571 code in it entered the analysis. Time zero (*t*_0_) is the date of the first cirrhosis diagnosis, the first code specific to cirrhosis in the record (ICD-10 K70.3, K71.7 or K74.3 to K74.6; ICD-9 571.2, 571.5 or 571.6), or the first K70, K74 or 571 code for the three patients without one, and is independent of AUD diagnosis status. The record starts on 1 January 2015, the lower bound on *t*_0_: a patient whose first cirrhosis code fell before that date enters with *t*_0_ in early 2015. Observed *t*_0_ runs from January 2015 to February 2023, and 99.8% of the registry enters by the end of 2020.

The study cohort comprised the 485 patients drawn by stratified sampling from the ICR^26^. The charts were drawn within strata of AUD diagnosis status at any time in the structured record, in roughly equal numbers from the 4,210 registry patients with an AUD diagnosis and the 10,196 without, and unequally across hospital types (Registry estimate). Extended Data Fig. 1 gives the flow of patients from the registry to the analyzed set.

### Clinical notes

For each patient in the study cohort, the notes were every note retrieved in a window of 18 months either side of that patient’s study-cohort index date. The study-cohort index date is the first recorded AUD diagnosis for the 231 patients sampled as having an AUD diagnosis whose record includes one and the first recorded cirrhosis diagnosis for the remaining 254. For patients without an AUD diagnosis, the hepatologists judged the cirrhosis diagnosis the point in the record most likely to include a discussion of alcohol. The study-cohort index date anchors the note window and is a different quantity from the first cirrhosis diagnosis of the registry analysis (*t*_0_).

The notes came from three hospital systems: an academic health system (224 patients), a safety-net hospital (57), and other contributing hospitals (204). Each patient in the study cohort was assigned to the hospital system that wrote most of that patient’s delivered notes, using the data provider’s report on the delivered notes.

### MedDx

MedDx reads each patient’s notes under the skill and returns one of six grades, which collapse to the three classes used throughout: AUD+, AUD− and indeterminate (Supplementary Table 1). It has three components, the skill, the harness and the engine.

#### The skill

The skill is an eight-block extraction and classification instruction, obtained by structured elicitation of hepatologist A’s own chart-review reasoning, and is printed in full in Supplementary Note 3. The eight blocks state four rule findings, each sufficient for AUD+, and require with every classification the rule findings, the note type or section cited and a verbatim quotation. The skill was completed before any MedDx output was compared with a ground-truth classification. Every chart was processed once under the skill, and the same skill was used for every hospital system.

#### The harness

The harness is the fixed program that runs the skill on the engine, one patient at a time. It passes the skill and the patient’s notes, concatenated into one input, to the engine and returns the classification in a fixed set of fields: one of the six grades, the rule findings, the note type or section, the supporting quotation and a confidence. When a patient’s notes exceed the engine’s input limit, the harness divides them into parts, classifies each part and keeps the strongest of the five grades from definitely no to definitely yes that any part received. Not enough data stands only when every part received it. The quotation, note type or section and confidence are those of the part with the strongest grade, and the rule findings are pooled across parts. Whether each quotation is verbatim in the notes was assessed afterwards (Quotation check).

#### The engine

The engine is the open-weight language model that reads the notes under the skill. Eleven engines were evaluated on the same 485 charts under the same skill and harness, and their repositories, revisions, release dates, sizes and numeric formats are in Supplementary Table 14. The eleven span eight developers, parameter counts from 21.5B to 753B and release dates from November 2024 to July 2026. Each was the current instruction-tuned release of its line at the time of evaluation. Qwen3.6-27B is the primary engine (Engines and skill variations).

### Ground truth

Chart review was performed by four hepatologists at Indiana University School of Medicine. Hepatologists A and B are hepatology fellows, hepatologist C is Associate Professor of Medicine and hepatologist D is Professor of Medicine. Hepatologist A read every chart, hepatologist B read a random subsample, hepatologist C gave second readings and hepatologist D adjudicated. All four read blind to all MedDx output.

#### The classification rule

The skill states the same classification rule the hepatologists used, having been elicited from the hepatologist who wrote their abstraction form, so agreement between MedDx and a hepatologist measures how reliably one rule is executed on a chart. Each of four rule findings is sufficient for definitely yes: alcohol-associated cirrhosis named as such, an explicit diagnosis of alcohol use disorder, dependence or abuse, an AUDIT score of 8 or more or a CAGE score of 2 or more, and documented withdrawal or its management. Current sobriety leaves the grade unchanged. Alcohol-attributed disease other than cirrhosis reaches probably yes on its own. Where a structured field and the narrative conflict, the narrative governs, and a structured field stands alone only where no narrative mentions alcohol. The hepatologists’ abstraction form searched each chart under eight headings: diagnosis and problem-list terms, drinking quantity and pattern, the AUDIT and CAGE instruments, withdrawal and its management, alcohol-attributed disease, treatment and recovery, supportive care, and social and legal circumstances.

#### Second reading and adjudication

Hepatologist A read every chart in the study cohort and returned one of the six grades. A chart went on to a second reading by hepatologist C for either of two reasons (Extended Data Fig. 2). The first was disagreement with MedDx. MedDx was run with three engines, Qwen3.6-27B, GPT-OSS-120B and GLM-5.2, and a chart went to a second reading when its classification under any one of the three differed from hepatologist A’s at the three-class level, which selected 57 charts. The second was disagreement between two hepatologists. Hepatologist B read a random subsample of 186 charts, blind to hepatologist A’s classifications, and took no part in adjudication. Hepatologists A and B differed on 15 charts at the three-class level. Seven were already among the 57 selected for the first reason, and the remaining 8 went to a second reading for this reason alone.

Hepatologist C read 65 charts: 50 for the first reason alone, 8 for the second alone and 7 for both. Hepatologist D adjudicated the 36 on which hepatologist C and hepatologist A differed in class. The ground truth is hepatologist D’s classification on the charts he read, hepatologist C’s on the other charts with a second reading and hepatologist A’s on the charts with a single reading, whichever reason led to the second reading. Of the 485 classifications, 420 are hepatologist A’s, 29 are hepatologist C’s and 36 are hepatologist D’s. The share of charts each hepatologist classified and the agreement between hepatologists are in Supplementary Table 3.

#### Verification-independent ground truth

The first reason for a second reading depends on MedDx output, so charts were verified at different intensities according to what MedDx returned (differential verification^45^). A verification-independent ground truth was therefore built from the second reason alone, over the 186 charts hepatologist B read. It takes hepatologist A’s classification where hepatologists A and B agree and the classification of hepatologist C or D where they differ, so its charts were selected without reference to MedDx. Of the 186, 161 have an AUD+ or AUD− ground truth, 96 AUD+ and 65 AUD−, and the remaining 25 are indeterminate and contribute no scored comparison. Registry-weighted specificity against it is reported in the Results, under weights formed over the charts that contribute to it.

### Registry estimate

Structured diagnoses were grouped by code set. An AUD diagnosis is an ICD code for AUD (F10.x, ICD-9 303.x or 305.0x). An alcohol-related diagnosis other than AUD is any alcohol-related code that is not an AUD code, including alcohol-associated liver disease (K70, ICD-9 571.0–571.3). No alcohol-related diagnosis is the absence of any code in the set. At the first cirrhosis diagnosis (t0), each patient falls into exactly one of three strata defined by these categories. ICD-9 codes are included because the record spans the October 2015 transition, and a K70-only definition of alcohol-associated liver disease is confounded with calendar time. Code lists are provided in Supplementary Table 15.

The registry estimate of the number of patients with AUD is the ground truth, post-stratified to the registry. The 423 reviewed charts with an AUD+ or AUD− ground truth are included, and the 62 indeterminate charts are described and carry no weight (Extended Data Fig. 3). Every registry patient carries a hospital type, determined by the sites of the patient’s inpatient and emergency visits: safety-net hospital if any visit was there; academic health system if any visit was there; otherwise, other hospitals. Every registry patient also carries a whole-record diagnosis status: an AUD diagnosis at any time, an alcohol-related diagnosis other than AUD, or neither. The nine cells of hospital type and diagnosis status contain the 14,406 registry patients and the 423 reviewed charts with an AUD+ or AUD− ground truth (Extended Data Fig. 4; Supplementary Table 5).

Within each cell, the share with AUD is the share of its AUD+ or AUD− charts whose ground truth is AUD+, and the cell’s patients with AUD are that share multiplied by its registry patients. The registry estimate is the sum over the nine cells, totaling 6,223 patients. Equivalently, each AUD+ or AUD− chart carries the weight of the registry estimate, the registry patients of its hospital type and whole-record diagnosis status, over the AUD+ or AUD− charts in that cell that contribute to the estimate^46^. Any count is then the weighted number of AUD+ charts: the count within a stratum at *t*_0_, a hospital type, or a patient group is the weighted number of AUD+ charts among the reviewed charts in that stratum, type, or group. The same weights, applied to MedDx’s classifications of the same charts, yield the MedDx projection in Fig. 2b.

Intervals are percentile bootstraps over the AUD+ or AUD− charts, resampled within the cells, with 2,000 resamples. Weights are set by cell, and a stratum at *t*_0_ cuts across cells, so a stratum’s interval can exceed its registry size. The upper bound of the other alcohol-related diagnosis stratum exceeds its registry size, and that of the AUD diagnosis stratum does as well, so the AUD diagnosis stratum’s share is reported as a point.

#### Sensitivity analyses and assumptions

Five specifications vary the registry estimate (Supplementary Table 16). The indeterminate charts were counted as AUD− (lower bound) and AUD+ (upper bound), with a tipping-point series between the lower bound and the primary estimate. The cells were reduced to hospital type and AUD diagnosis alone, resulting in six cells. The hospital type of a reviewed chart was taken from the source of its notes rather than its visits, while the registry retained the visit-based type.

The registry estimate rests on two assumptions. First, within a cell defined by hospital type and whole-record diagnosis status, entry into the study cohort and the AUD+ or AUD− ground truth are independent of AUD status. Second, the ground truth measures the target condition, defined as lifetime AUD, regardless of whether AUD is present at t0.

### Agreement and accuracy

The target condition is chart-documented lifetime AUD. Performance against the ground truth was measured by agreement under the primary definition below and, on the three-class collapse, by sensitivity, specificity, positive predictive value (PPV) and negative predictive value (NPV). Patients whose ground-truth classification is indeterminate are excluded from sensitivity and specificity. An indeterminate output on a patient with an AUD+ or AUD− ground truth stays in the denominator, so an indeterminate MedDx output on a ground-truth-positive patient counts as a patient MedDx did not identify. The intervals are described under Statistical analysis.

The primary agreement statistic is unweighted κ^47^ on the three classes, AUD+, AUD− and indeterminate. The three classes are the decision the classification supports, and indeterminate is an output state with no position between the two directions, so the classes carry no distance weights. A chart is excluded when either of the two classifications compared is not enough data, so the primary denominator is 428 of the 485 charts and the indeterminate class of the primary κ contains uncertain alone. Counting not enough data in that class over all 485 charts gives 0.93 (Supplementary Table 17), and agreement within each of MedDx’s six grades is in Supplementary Table 2. The three-class collapse used for the descriptive counts in Fig. 1c pools uncertain and not enough data, on a different denominator from that of the κ. A weighted κ on the full six-grade scale requires a position for not enough data, which has no natural one. Across the six positions it can occupy, the same comparison gives 0.89 to 0.92.

Six definitions are reported beside the primary one in Supplementary Table 17, for MedDx against the ground truth and for both comparisons against hepatologist B. They are the same three classes with not enough data kept as indeterminate, with linear weights^48^ and with Gwet’s AC1^49^, the two directional classes restricted to charts on which both classifications were directional, the five classes that remain when not enough data is dropped and the scale is ordinal, and the six-grade metric under both orderings. Weights on the six-grade κ are linear (Supplementary Table 17). The conclusions of the agreement analysis hold under all six. The two comparisons against hepatologist B use first-pass classifications, before adjudication. Agreement between hepatologists A and B measures the reproducibility of the reading, and MedDx against hepatologist B on the same charts is reported beside it under the same definitions (Supplementary Table 17).

Estimates are reported on the study cohort and weighted to the registry with the weights of the registry estimate (Registry estimate). Sensitivity, specificity and Youden’s J^29^ are reported unweighted and weighted, and the weighted form describes the registry. The share of charts with a directional classification, and its complement the indeterminate rate, are reported beside accuracy on the same denominator. Predictive values are reported for the registry. The positive predictive value is the weighted share of the charts classified AUD+ whose ground truth is AUD+, and the negative predictive value the weighted share of those classified AUD− whose ground truth is AUD−, with the bootstrap of the registry estimate. Fig. 1f draws both against prevalence from the weighted rates of AUD+ and AUD− classification among ground-truth-positive and ground-truth-negative charts, an indeterminate output counting as neither.

### Comparators

The comparator set spans both data sources. Two read the structured record, a code rule and a supervised model, and two read the same notes MedDx read, both supervised models. Each was scored against the same ground truth on the same AUD+ or AUD− charts. Variants of each and full metrics are in Supplementary Tables 18 and 19.

The rule comparator flags any alcohol-related diagnosis, a code set wider than F10.x and given in Supplementary Table 15. A variant adds alcohol-associated liver disease to the AUD diagnosis. The supervised comparator is a gradient-boosted tree over age, sex, race, ethnicity, the 19 Charlson conditions, the comorbidity index, encounter and note counts, and two alcohol-code flags. Its features exclude hospital system. It was fitted with five-fold outer and three-fold inner cross-validation and is reported out of fold, both with and without the AUD diagnosis among its features. The version without the AUD diagnosis is the principal supervised structured comparator, because the patients of interest are those the diagnosis does not identify.

Two comparators read the same notes as MedDx. The first is a TF-IDF logistic regression: an L2-regularized logistic regression over word and bigram counts with sublinear term frequency and inverse document frequency, fitted on each patient’s concatenated notes. The second is Bio_ClinicalBERT^30^, a BERT encoder pretrained on clinical notes and fine-tuned for this classification. A patient’s concatenated notes exceed the encoder’s 512-token input, so they were divided into non-overlapping 512-token windows, each of which inherits the patient’s ground-truth classification. Training used at most 32 windows per patient, taken at even spacing through the notes, which limits the weight of a long chart in the loss. At inference every window was scored and the patient’s score is the mean over its windows.

Both text comparators used the same nested cross-validation as the structured model. Every fold, outer and inner, was drawn over patients and the windows were cut after the split, so all of a patient’s text lies on one side of every split. The operating threshold for every supervised comparator is the quantile of the training fold’s own score distribution taken at that training fold’s ground-truth-positive prevalence, so the threshold uses training-fold classifications only.

#### Transfer across hospital systems

The gradient-boosted model with the AUD diagnosis among its inputs and the TF-IDF logistic regression were also fitted on the AUD+ or AUD− charts of the academic health system alone (191 charts), with the same features, grids and nested cross-validation as above. Each was then applied unchanged and scored on the safety-net hospital (56 charts) and the other hospitals (176). The classification threshold was the quantile of the out-of-fold scores at the academic health system’s ground-truth prevalence, carried unchanged. Sensitivity and specificity by system have Wilson 95% CI. The academic-system values are the 5-fold out-of-fold estimates on the training charts (Fig. 3e; Supplementary Table 8).

### AUD-specific medication

The outcome is the first recorded receipt, on or after the first cirrhosis diagnosis (*t*_0_), of any of the three medications approved for AUD in the United States: naltrexone (standard concept 1714319), acamprosate (19043959) or disulfiram (735850), with descendants. Methylnaltrexone (909841), a peripheral opioid antagonist, was excluded, as were the morphine–naltrexone and bupropion–naltrexone combination products, which are not AUD treatment. Gabapentin, baclofen and topiramate are not counted, because their recorded use before *t*_0_ does not follow the alcohol-related diagnoses (Supplementary Fig. 4). A patient with receipt before *t*_0_ is a prior user, counted as treated at *t*_0_ and outside the time-to-event analysis. Every other patient is a new user, followed on a time axis from *t*_0_.

Among patients with AUD in a stratum at *t*_0_, the share on an AUD-specific drug by year t is the stratum’s prior users plus its new users who first received one within t years of *t*_0_, over the registry estimate of patients with AUD in the stratum. The new users treated by year t are the stratum’s new users times the Aalen–Johansen cumulative incidence of a first AUD-specific drug, with death as a competing event, from *t*_0_ to the first of the drug, death, or the end of the record on 10 July 2023 (Extended Data Fig. 7a). The prior users and the study-cohort charts of each stratum are in Extended Data Fig. 7b,c. The gap between strata is the difference in shares, at *t*_0_ and at five years, and the ratio is the ratio of the five-year shares (Extended Data Table 2). Intervals are percentile bootstraps over 500 paired resamples, the AUD+ or AUD− charts within cells for the denominator and the stratum’s new users for the cumulative incidence.

Deaths are recorded whether or not a patient is still seen, so follow-up was censored at the end of the record. A patient no longer seen counts as untreated and at risk, which lowers the shares. In the sensitivity analysis follow-up was censored at each patient’s last record, which moves the shares in the other direction (Extended Data Table 2). The shares count every recipient of an AUD-specific drug as a patient with AUD, the third assumption of the registry-scale analysis, on the ground that a patient with cirrhosis who receives one of these three drugs has the disorder.

### Patient groups and hospital systems

Performance was computed within each hospital system with the measures above, with the primary κ for agreement (Agreement and accuracy). The paired gain reported for a hospital system is MedDx’s sensitivity there minus the AUD diagnosis’s on the same charts, in percentage points, with a percentile bootstrap interval over the patients of that system. Agreement was also computed within three calendar periods of the first cirrhosis diagnosis, 2015–16, 2017–18 and 2019 onward (Supplementary Fig. 1).

Differences between patient groups were assessed at two scales, the reviewed charts and the registry, each with the variables recorded for every patient at that scale. At chart scale, six variables supply 18 groups: hospital system, sex, race, age group, urban–rural classification and neighborhood deprivation quintile. At registry scale, five variables supply 15: age group, sex, race, Charlson index and neighborhood deprivation quintile. Hospital system and urban–rural classification were linked for the reviewed charts alone, and the Charlson index is reported at registry scale only. Age is grouped as under 50, 50 to 64 and 65 and over. A patient with no recorded value of a variable is in none of that variable’s groups.

Neighborhood deprivation is the Social Deprivation Index of the Robert Graham Center, built on the 2015–2019 American Community Survey and published for each ZIP code tabulation area. The five-digit ZIP recorded for each patient was matched to the tabulation area of the same five digits. The published score is a population-weighted national percentile, so each 20-point band contains one fifth of the United States population and defines a quintile without reference to these data. The index was linked for 14,207 of the 14,406 registry patients and 481 of the 485 reviewed charts. Comorbidity burden used the Charlson index in the Glasheen specification^50^ with the National Cancer Institute code lists^51^, applying the hierarchy rules under which a severe condition supersedes its milder form and requiring co-occurring HIV and an opportunistic infection for the AIDS component. The two liver components were excluded because every registry patient has cirrhosis.

At chart scale, the quantity is the difference in sensitivity and in specificity between a group and all other groups of its variable, in percentage points, computed for the AUD diagnosis and for each model on the same charts, with zero as no difference and a 4,000-sample bootstrap. The reference band is 5 points, the reporting threshold, and the intervals are given beside it.

Two summaries of the 18 chart-scale differences are reported beside the counts of groups outside the band, for the AUD diagnosis and for each model. The span on a metric is the largest of the 18 differences minus the smallest. The same 18 groups are also placed as operating points in a plane whose axes are the group’s false positive rate and its sensitivity, both in percentage points, and the area of the convex hull of the 18 points is reported in square points. The same estimator on the same 18 groups was applied to the 11 engines of Fig. 4b and to the skill with its four variations of Extended Data Fig. 5.

At registry scale, the quantity is the sensitivity of the AUD diagnosis by *t*_0_ among registry patients estimated to have AUD, which uses the weights of the registry estimate (Registry estimate) within the group. For a group, the estimated number of patients with AUD is the weighted number of AUD+ charts among its reviewed charts. Sensitivity is the weighted share of them in the stratum that carries an AUD diagnosis at *t*_0_, and the remainder are the patients with AUD in the group who carry no AUD diagnosis at *t*_0_. The reviewed charts alone cannot supply this quantity because they were drawn stratified on diagnosis status. Confidence intervals are the percentile bootstrap of the registry estimate, 2,000 resamples of the AUD+ or AUD− charts within cells. Sensitivity in a group is also expressed as a ratio to the same quantity in all other groups of that variable, with one as no difference. Intervals for the ratio are 2,000-sample percentile bootstrap intervals over the AUD+ or AUD− charts, resampled within the cells of the weights. The band on that axis is the four-fifths rule, 0.8 to 1.25, whose edges multiply to one.

#### Indeterminate rate by group

The rate of indeterminate classification was stratified separately from accuracy, on a denominator of every chart in the group, including the charts whose ground-truth classification is itself indeterminate. Seven variables were used, each taken from the structured record or from the patient’s ZIP: hospital system, sex, race, age group, decompensation at the first cirrhosis diagnosis, first cirrhosis diagnosis before or from March 2020, and urban–rural classification. Both indeterminate grades count for MedDx, matching the single indeterminate class of the ground truth, and the same quantity was computed for the hepatologists on the same charts. Intervals are Wilson intervals. One of the 485 charts has no urban–rural classification, so each rate is computed on the charts assigned to a level of its variable. Every group is in Supplementary Table 9.

### Engines and skill variations

Qwen3.6-27B is the primary engine because it agreed most closely with the ground truth on these 485 charts. The three engines with the highest agreement are marked as recommended in Fig. 4b, and GPT-OSS-20B and every engine ranked below it are marked as not recommended. Agreement between every pair of engines was computed under the primary definition, with the ground truth entered as one more row (Fig. 4c), and rows and columns keep the order of Fig. 4b. Patients were resampled 2,000 times and every engine’s agreement recomputed in each resample, recording how often each engine came first and the 2.5th to 97.5th percentile of its rank (Supplementary Table 13).

The skill was run at temperature 0. Four variations were run: the same skill at temperature 0.3 and at temperature 0.7, and two paraphrases of its text, paraphrased skills A and B, each at temperature 0. A paraphrase rewrites the wording of the eight blocks and leaves the classification rules, the six-grade scale and the required output fields unchanged. Both are released with the analysis code (Supplementary Note 3). Extended Data Fig. 5 shows the absolute agreement of the skill and of each variation, with a percentile bootstrap over patients. Fig. 4e shows the stability of the reading on the 186 charts hepatologist B read. For each variation it gives the κ between the classification under that variation and under the skill at temperature 0, beside the same statistic for hepatologist A against hepatologist B and for MedDx against each of them, each with a percentile bootstrap interval over patients. The paired change from the skill at temperature 0 uses a paired bootstrap over the charts both conditions placed.

### Quotation check

Each classification carries a supporting quotation that MedDx drew from the notes. Each quotation was compared with the notes MedDx read, ignoring differences in letter case, punctuation and spacing. A quotation is verbatim when it matches one passage of the notes word for word, or when it joins non-adjacent passages with an ellipsis and every passage matches. The remaining quotations are not verbatim and were separated by whether every word occurs in the patient’s notes. The check shows that a quotation is in the notes, not that it supports the classification. Where MedDx returned an explanatory statement in place of a quotation, the classification is reported separately.

#### Note types or sections and rule findings

The harness returns two further fields with every classification. The rule findings are the enumerated list the skill defines (Supplementary Note 3, section 3.1), and a classification returning none of them is recorded as having no rule finding. The note type or section is MedDx’s own free-text report of where in the notes its evidence lay. The skill gives examples and leaves the set open. The figures show the ten most cited (Fig. 4g; Extended Data Fig. 6b) and the eight most cited among ground-truth-positive patients (Extended Data Fig. 6a). A classification may cite more than one note type or section, so citations exceed classifications, and in Extended Data Fig. 6a a patient is counted in every pair of rule finding and note type or section its classification carries. A classification carrying none of the four rule findings is drawn as other evidence, outside the rules. The third rule finding, a positive AUDIT or CAGE score, was carried by no patient. The 62 ground-truth-indeterminate patients are omitted from these panels.

### Statistical analysis

Unless stated otherwise, the following apply. Bootstrap confidence intervals used the percentile method with 2,000 resamples. The chart-scale patient-group differences and the agreement definitions used 4,000 resamples, and the treatment shares used 500 paired resamples. Intervals for binomial proportions are Wilson intervals^52^, and the share of quotations verbatim in the chart has a Clopper–Pearson exact interval^53^. Registry-scale quantities carry percentile bootstrap intervals. Estimated counts, shares and differences are each rounded from the unrounded value, so printed components can differ from a printed total by one in the last digit. The symmetry of the disagreements in Fig. 1c was tested with Bowker’s test. Paired comparisons of correctness used the discordant pairs only^54^: an exact binomial test where there are fewer than 25 of them, and a continuity-corrected McNemar test otherwise. All analyses were conducted in Python 3.12.

## Ethics

The Institutional Review Board of Indiana University waived ethical approval for this work (exempt determination, protocol 29101, exemption category 4(iii)). As secondary research on existing health records, it did not require informed consent.

## Data availability

The clinical notes and patient-level data used in this study contain protected health information and cannot be shared publicly. Summary statistics and de-identified analysis outputs are available in the Supplementary Information. Access to source data may be requested through the Indiana Network for Patient Care data governance process.

## Code availability

The analysis code, from the hepatologists’ classifications and the per-patient model classifications onward (the ground truth, the estimators, the registry estimate and the figure and table producers), is at https://github.com/Su-informatics-lab/aud. The skill is printed in full in Supplementary Note 3 and released under CC-BY 4.0 as a standalone Agent Skill in the same repository.

## Author contributions

H.W. and J.S. conceived the study. H.W. developed MedDx, performed the analyses and wrote the manuscript. Y.C. verified the data and the analyses. C.X. and H.L. contributed to the formal analysis. D.P. managed the registry data. M.A., A.P.D. and S.G. reviewed the charts and contributed to the clinical interpretation. S.L. contributed to the clinical interpretation. W.T. supervised the statistical analysis. J.S. supervised the study.

## AI Use Declaration

Large language models are the subject of this study: the skill, the harness and the engines of MedDx and their evaluation are described in the Methods. Large language model assistants (Claude, Anthropic; Codex, OpenAI) were used for code development, figure preparation and language editing. The authors reviewed and verified all AI-assisted output and take full responsibility for the content of this article.

## Extended Data

**Extended Data Fig. 1.**
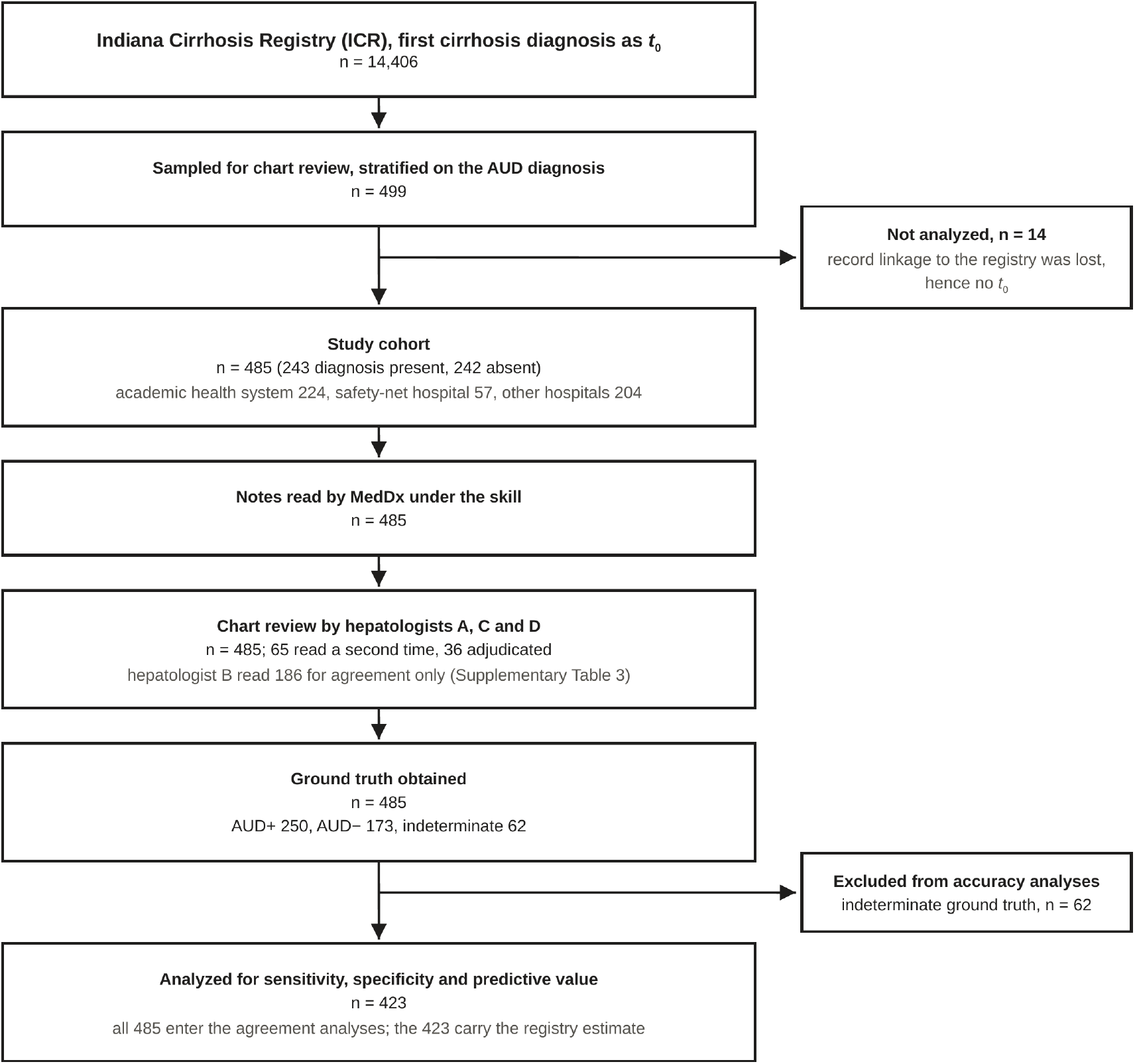
Flow of patients from the ICR to the analyzed set. Charts were sampled stratified on whether the structured record carried an AUD diagnosis at any time, which oversamples the diagnosis-present group relative to the registry and is the reason registry-scale estimates are weighted to the registry (Methods). MedDx read the notes of every patient in the study cohort and every one received hepatologist chart review. The charts were sampled from an earlier version of the registry data than the one analyzed, and patient record linkage was revised between the two. Record linkage to the registry was lost for 14 of the 499 sampled patients, who are not analyzed. Of the 243 patients sampled as having an AUD diagnosis, 231 carry one in the structured record analyzed, as do 2 of the 242 sampled as having none (233 in all); that record defines the AUD diagnosis in every analysis. Patients whose ground truth was indeterminate, meaning the hepatologist judged the record insufficient or was unable to decide, have no class to score against and are excluded from the accuracy analyses. They are described alongside the rest of the study cohort in Extended Data Table 1.

**Extended Data Fig. 2.**
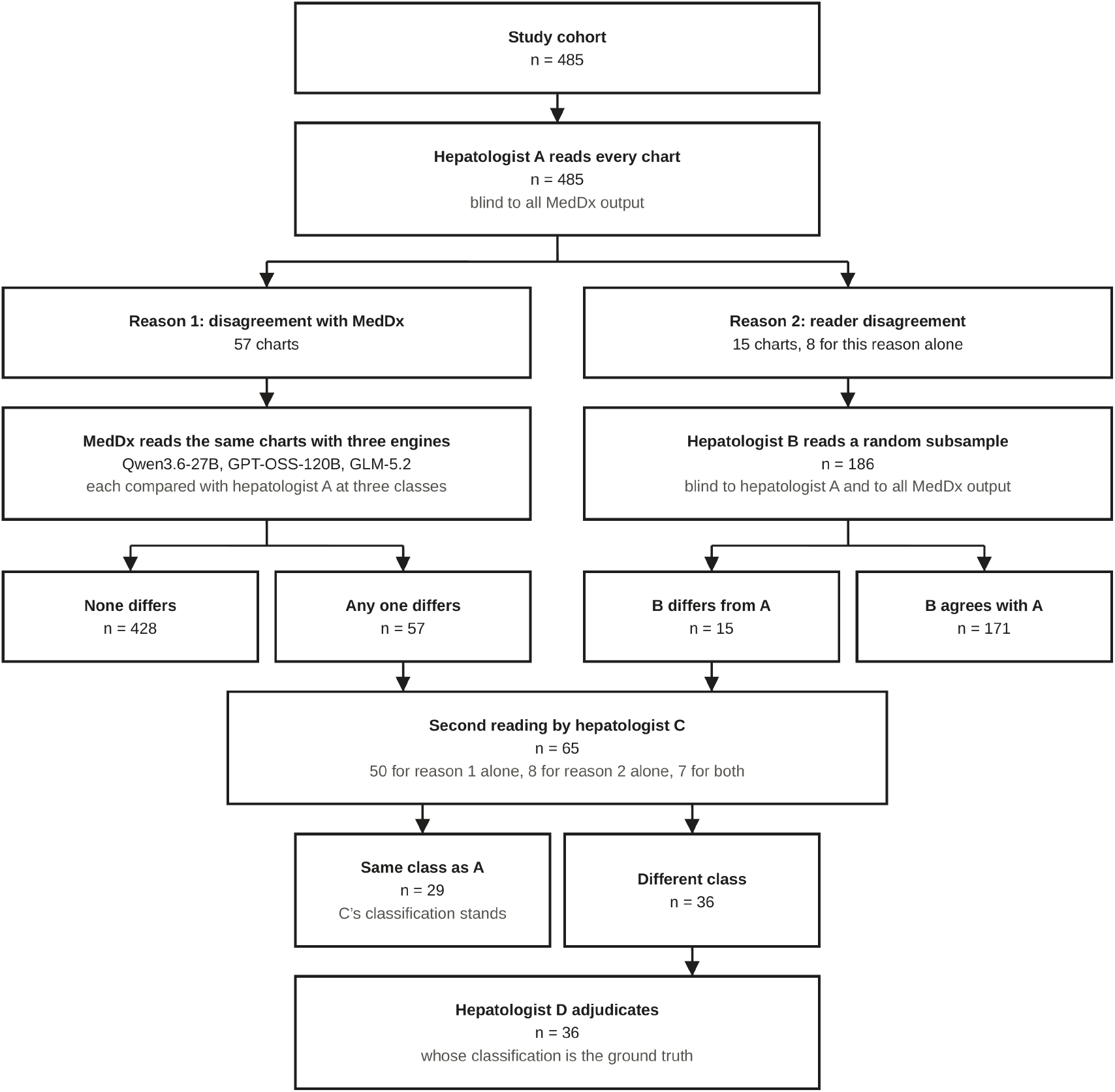
The ground truth: one procedure, two reasons for a second reading. The figure expands the box Ground truth obtained in Extended Data Fig. 1. All four hepatologists read blind to MedDx output. Hepatologist D’s classification is the ground truth wherever D read the chart, hepatologist C’s stands where D did not, and hepatologist A’s stands where neither read the chart. Of the 485 classifications, 420 are A’s, 29 are C’s and 36 are D’s. Hepatologist B’s classifications are not part of the ground truth and enter only the agreement analyses (Fig. 4e).

**Extended Data Fig. 3.**
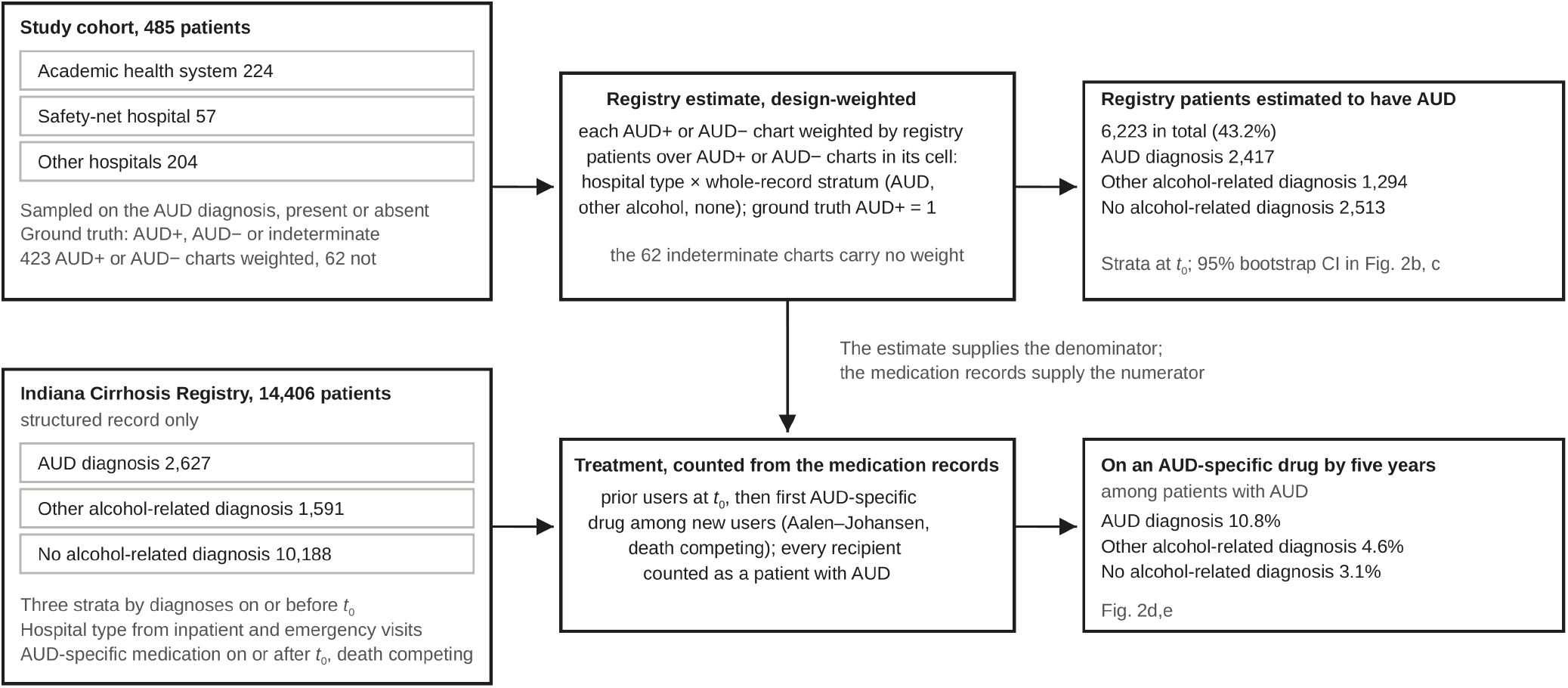
How the study cohort reaches the registry. The study cohort supplies the ground truth. Each of the 423 charts with an AUD+ or AUD− ground truth is weighted to the registry patients of its cell, hospital type by whole-record diagnosis status, and the 62 indeterminate charts carry no weight (Methods). The weighted sum is the registry estimate, 6,223 patients with AUD, which is the denominator for the treatment shares; the medication records supply the numerator. Hospital type is a weighting cell, whereas the hospital system that wrote a patient’s notes is the variable on which the classifiers are compared (Fig. 3c; Supplementary Table 7).

**Extended Data Fig. 4.**
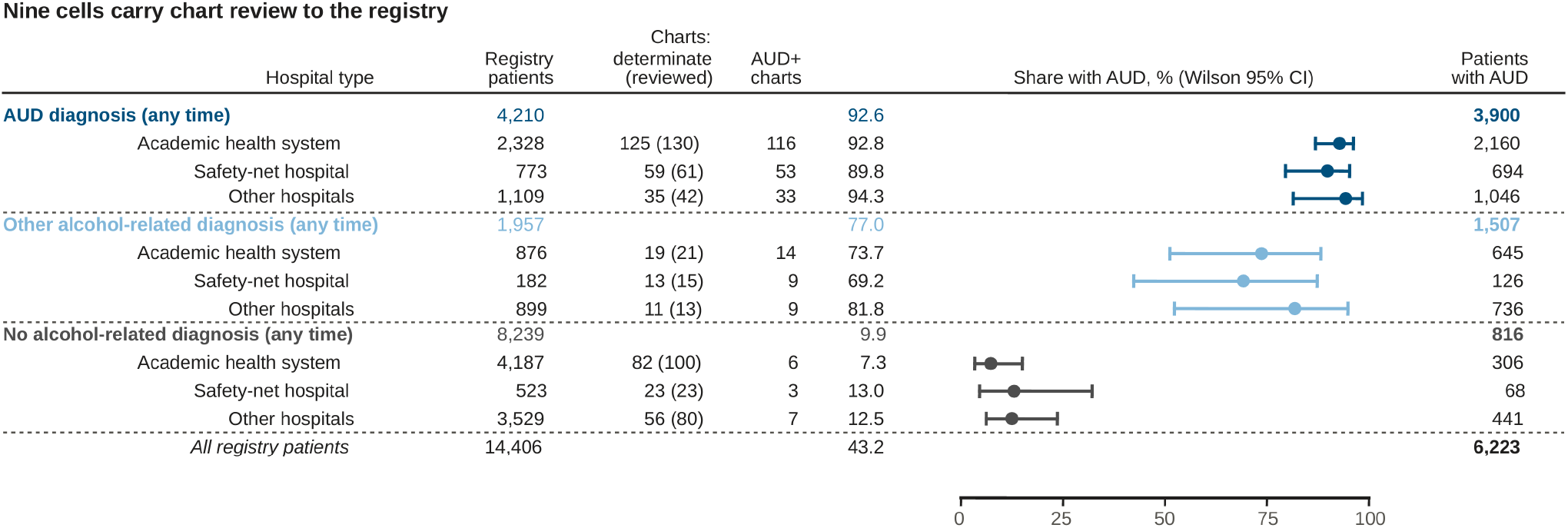
The nine cells of the registry estimate. Hospital type, from the sites of a patient’s inpatient and emergency visits, by whole-record diagnosis status: an AUD diagnosis at any time, an alcohol-related diagnosis other than AUD, or neither. Each cell gives its registry patients, its reviewed charts with an AUD+ or AUD− ground truth (of all its reviewed charts), the ground-truth-positive charts among them, the share with AUD with a Wilson 95% CI, and the registry patients with AUD that share implies. The stratum lines sum the cells, and the total is the registry estimate of Fig. 2b. The same cells are tabulated in Supplementary Table 5.

**Extended Data Fig. 5.**
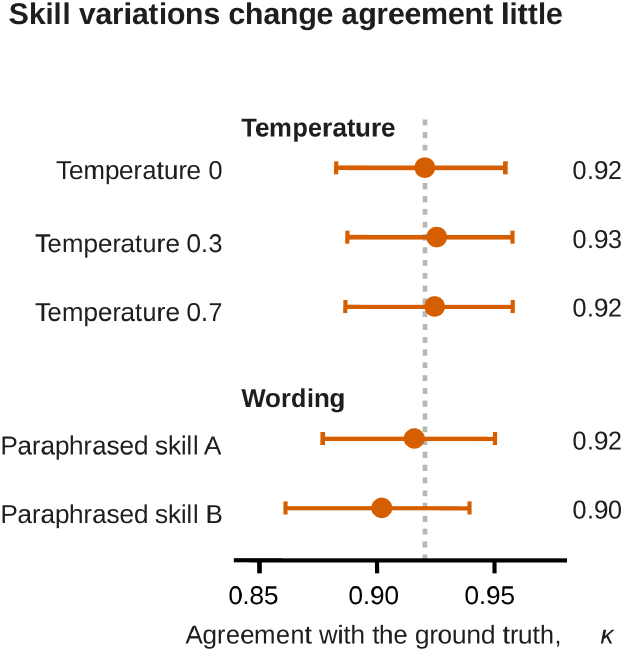
Agreement with the ground truth under the skill and four variations. Two variations change the decoding temperature and two the wording of the skill; each is run on the 485 charts of the study cohort. Agreement is unweighted Cohen’s κ on three classes, a chart being dropped when either classifier returned not enough data, with 2,000-sample percentile bootstrap intervals over patients. The dotted line is the skill’s value at temperature 0. The paired change of each variation from it is in Results.

**Extended Data Fig. 6.**
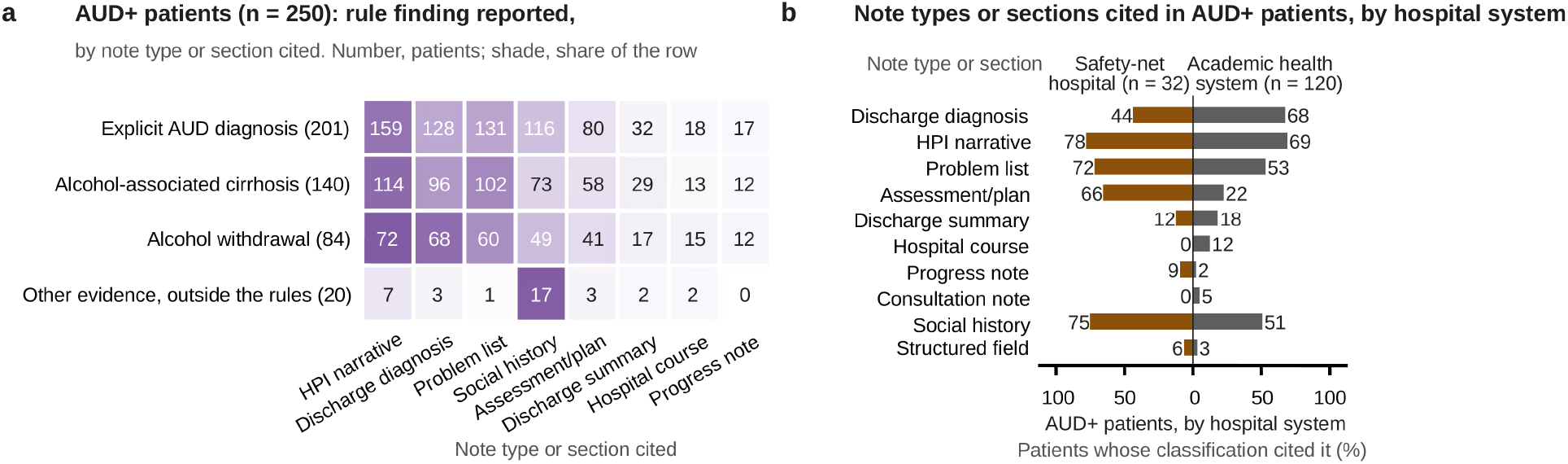
Where MedDx found its evidence in the chart. **a**, The 250 ground-truth-positive patients by rule finding (rows) and note type or section cited (columns), eight most cited; number: patients, shade: share of the row. Rule findings are named as in the skill, and an explicit AUD diagnosis is one written in the notes. Of the 20 on other evidence, outside the rules, 17 cited the social history. **b,** The same ten note types or sections as in Fig. 4g, among the AUD+ patients by hospital system: the share at the safety-net hospital (brown, *n* = 32) and the academic health system (gray, *n* = 120).

**Extended Data Fig. 7.**
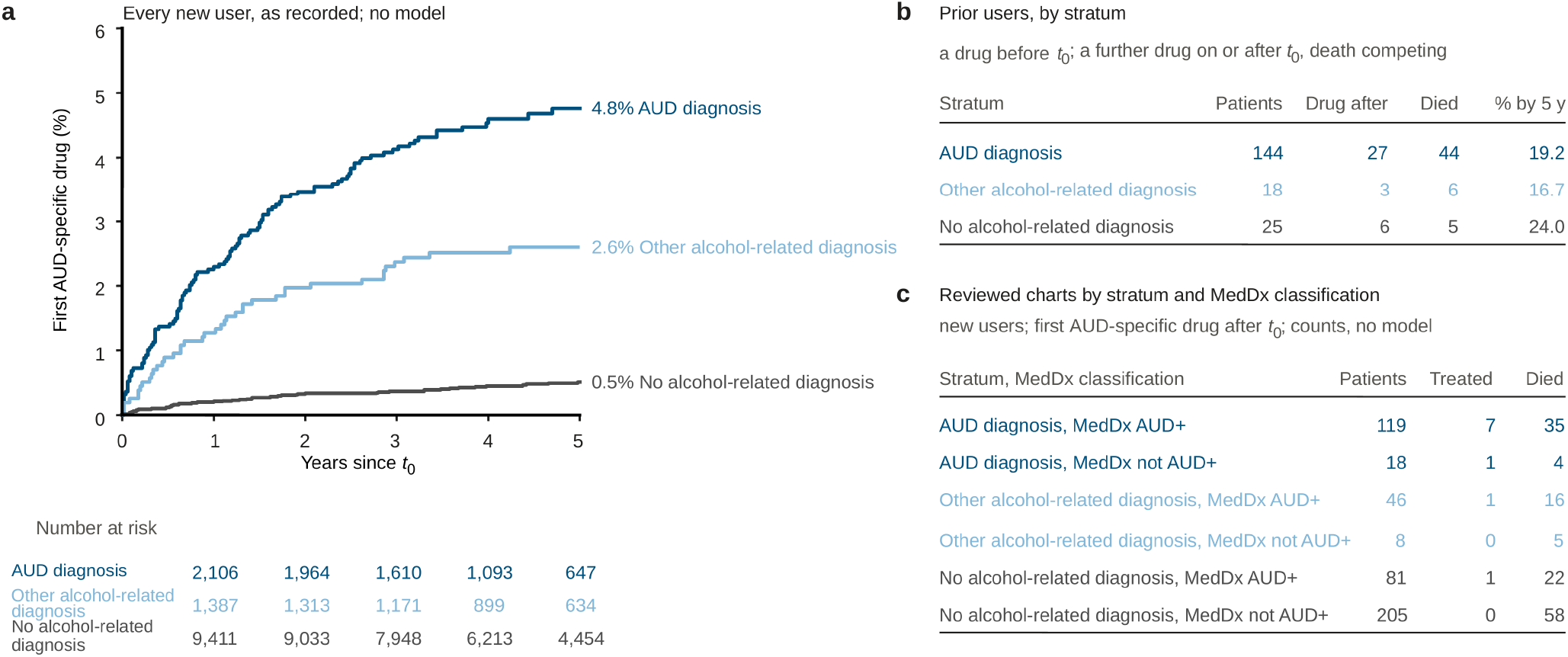
AUD-specific medication after *t*_0_ in the registry, as recorded. **a**, Cumulative incidence of a first naltrexone, acamprosate or disulfiram after *t*_0_ among every new user of each stratum, with death as a competing event (Aalen–Johansen), and numbers at risk below. The value at the right is at five years. These curves include the patients in each stratum who do not have AUD. **b**, The 187 prior users, patients with an AUD-specific drug recorded before *t*_0_ and therefore outside a: by stratum, how many had a further drug recorded on or after *t*_0_, how many died, and the cumulative incidence of a further drug by five years with death competing. **c**, The study-cohort charts of every stratum who were new users, by MedDx classification: patients, patients with a first AUD-specific drug after *t*_0_, and deaths, as recorded.

**Extended Data Table 1.** Characteristics of the study cohort. Continuous variables are mean (SD) with median [minimum, maximum] beneath; categorical variables are n (%) within column. Columns are the sampling strata: the reviewed charts were drawn stratified on whether the structured record carried an AUD diagnosis, in roughly equal numbers, so the overall column describes the study cohort and not the registry. Registry estimates are given in the main text.

|  | **AUD diagnosis absent (*n* = 242)** | **AUD diagnosis present (*n* = 243)** | **Overall (*n* = 485)** |
|---|---|---|---|
| Age at the study-cohort index date, mean (SD) | 63.0 (11.6) | 54.4 (11.3) | 58.7 (12.2) |
| Median [min, max] | 62 [24, ≥90] | 56 [25, 79] | 59 [24, ≥90] |
| *Sex* | — | — | — |
| Man | 115 (47.5%) | 154 (63.4%) | 269 (55.5%) |
| Woman | 127 (52.5%) | 89 (36.6%) | 216 (44.5%) |
| *Race* | — | — | — |
| White | 213 (88.0%) | 207 (85.2%) | 420 (86.6%) |
| Black | 20 (8.3%) | 34 (14.0%) | 54 (11.1%) |
| Other | 9 (3.7%) | 2 (0.8%) | 11 (2.3%) |
| *Ethnicity* | — | — | — |
| Hispanic | 7 (2.9%) | 11 (4.5%) | 18 (3.7%) |
| Other | 235 (97.1%) | 232 (95.5%) | 467 (96.3%) |
| *Hospital system* | — | — | — |
| Academic health system | 102 (42.1%) | 122 (50.2%) | 224 (46.2%) |
| Safety-net hospital | 32 (13.2%) | 25 (10.3%) | 57 (11.8%) |
| Other hospitals | 108 (44.6%) | 96 (39.5%) | 204 (42.1%) |
| Charlson comorbidity index, mean (SD) | 6.3 (3.1) | 5.5 (2.8) | 5.9 (3.0) |
| Median [min, max] | 6 [1, 16] | 5 [1, 16] | 5 [1, 16] |
| *Selected comorbidity* | — | — | — |
| Decompensated liver disease | 128 (52.9%) | 171 (70.4%) | 299 (61.6%) |
| Diabetes with complications | 91 (37.6%) | 39 (16.0%) | 130 (26.8%) |
| Renal disease | 69 (28.5%) | 46 (18.9%) | 115 (23.7%) |
| Congestive heart failure | 87 (36.0%) | 59 (24.3%) | 146 (30.1%) |
| Chronic pulmonary disease | 104 (43.0%) | 103 (42.4%) | 207 (42.7%) |
| Malignancy | 62 (25.6%) | 49 (20.2%) | 111 (22.9%) |
| Notes reviewed per patient, mean (SD) | 27.0 (32.9) | 28.6 (48.2) | 27.8 (41.2) |
| Median [min, max] | 16 [2, 269] | 16 [2, 431] | 16 [2, 431] |
| *Ground truth* | — | — | — |
| AUD+ | 38 (15.7%) | 212 (87.2%) | 250 (51.5%) |
| AUD− | 156 (64.5%) | 17 (7.0%) | 173 (35.7%) |
| Indeterminate | 48 (19.8%) | 14 (5.8%) | 62 (12.8%) |

**Extended Data Table 2.** AUD-specific medication after the cirrhosis diagnosis. Prior users are registry patients with naltrexone, acamprosate or disulfiram recorded before the cirrhosis diagnosis; new users are the rest, followed from that date to the first of these, death, or the end of the record on 10 July 2023, and the median is of that exit time. The share on a drug is the prior users plus the new users treated by that year (Aalen–Johansen cumulative incidence, death competing) over the registry estimate of patients with AUD in the stratum, every recipient counted as a patient with AUD; intervals are percentile bootstrap 95% CI (Methods). The last column censors each patient at the last dated record of any kind instead of on 10 July 2023. The cumulative incidence of a first drug by five years among all new users, with AUD or without, was 4.76%, 2.60% and 0.51% in the three strata (Extended Data Fig. 7a), and of death 30.5%, 24.5% and 22.1%.

| Stratum | Registry patients | Patients with AUD | Prior users | New users | First treatment | Deaths | Median follow-up, years | On a drug at the cirrhosis diagnosis, % (95% CI) | By 1 year | By 5 years | By 5 years, censored at last record |
|---|---|---|---|---|---|---|---|---|---|---|---|
| AUD diagnosis | 2,627 | 2,417 | 144 | 2,483 | 113 | 734 | 3.7 | 6.0 (5.3–6.8) | 8.3 (7.3–9.7) | 10.8 (9.4–12.7) | 12.0 |
| Other alcohol-related diagnosis | 1,591 | 1,294 | 18 | 1,573 | 41 | 410 | 4.5 | 1.4 (1.1–1.9) | 2.9 (2.1–4.4) | 4.6 (3.4–6.6) | 5.2 |
| No alcohol-related diagnosis | 10,188 | 2,513 | 25 | 10,163 | 53 | 2,491 | 4.6 | 1.0 (0.8–1.3) | 1.8 (1.4–2.6) | 3.1 (2.2–4.2) | 3.4 |
| Below the AUD diagnosis stratum, percentage points |  |  |  |  |  |  |  |  |  |  |  |
| Other alcohol-related diagnosis |  |  |  |  |  |  |  | 4.6 (3.6–5.6) |  | 6.3 (3.6–8.9) | 6.8 |
| No alcohol-related diagnosis |  |  |  |  |  |  |  | 5.0 (4.2–5.9) |  | 7.8 (5.7–9.9) | 8.5 |
| Ratio, AUD diagnosis stratum against |  |  |  |  |  |  |  |  |  |  |  |
| Other alcohol-related diagnosis |  |  |  |  |  |  |  |  |  | 2.4 (1.6–3.6) |  |
| No alcohol-related diagnosis |  |  |  |  |  |  |  |  |  | 3.5 (2.4–5.2) |  |

## Supplementary Information

### Supplementary Note 1. Second readings, sampling strata and the estimates in Fig. 1

A chart had a single reading only when MedDx agreed with hepatologist A under each of the three engines (Methods). Of the 423 charts with an AUD+ or AUD− ground truth, 365 had a single reading, and all 15 of MedDx’s errors and abstentions are among the 58 with a second reading. Of the 16 charts in which one of MedDx and hepatologist A was right and the other wrong, MedDx was correct in 10 (exact binomial *P* = 0.45; Fig. 1e). MedDx classified as AUD+ or AUD− 98.1% of the 423 patients with an AUD+ or AUD− ground truth, compared with 95.3% for hepatologist A.

The study cohort was drawn in balanced numbers on the AUD diagnosis, so charts with the diagnosis are over-represented relative to the registry. Among charts with an AUD diagnosis in the structured record the diagnosis identifies every ground-truth-positive patient, and among charts without one it identifies none, so its sensitivity and specificity depend on the mix of the two groups. The absence of a code is scored as a negative classification throughout. Weighted to the registry, the specificity of the AUD diagnosis is 96% (Fig. 1d). MedDx’s sensitivity is 99% among charts with the diagnosis (200 of 202) and 96% among charts without it (46 of 48), so weighting changes it little.

### Supplementary Note 2. The alcohol-associated cirrhosis finding and the other alcohol-related diagnosis stratum

The ground truth treats alcohol-associated cirrhosis named as such in the notes as sufficient for AUD+, and the K70 codes that place a patient in the other alcohol-related diagnosis stratum record the same condition in the structured record, so the two can agree without either adding information about alcohol use. The hepatologists’ review forms record a grade, a quotation and screening scores but not which finding decided the grade, so this check uses the rule findings MedDx reported for the same charts. On 29 of the 423 AUD+ or AUD− charts alcohol-associated cirrhosis was the only rule finding MedDx reported, 28 of them AUD+ in the ground truth; the 29 are 9 in the AUD diagnosis stratum, 8 in the other alcohol-related diagnosis stratum and 12 in the no alcohol-related diagnosis stratum.

Setting those 29 charts to indeterminate, so that they carry no weight, and leaving every other chart as it is gives a registry estimate of 6,044 patients with AUD (95% CI 5,518–6,559) against 6,223: 2,424 (92.3%) in the AUD diagnosis stratum, 1,129 (71.0%) in the other alcohol-related diagnosis stratum and 2,491 (24.5%) in the no alcohol-related diagnosis stratum. The shares on an AUD-specific drug by five years become 10.8% (9.2–12.6), 5.2% (3.5–7.9) and 3.1% (2.3–4.3), against 10.8%, 4.6% and 3.1%. Scoring the 29 charts as AUD− instead, the extreme case, puts the other alcohol-related diagnosis stratum at 61.1%, 972 patients, treated at 6.1%. The ordering of the three strata in prevalence and in treatment is the same in both scenarios.

### Supplementary Note 3. The skill and its extraction schema

The eight blocks are reproduced below verbatim, as the text every engine was given. The worked examples in its section 7 are mock examples written for the skill and contain no text from any patient’s chart. They are released as a standalone Agent Skill under CC-BY 4.0 (Code availability); the two paraphrased skills used as skill variations (Extended Data Fig. 5) are provided with the analysis code. The schema requires the engine to return, for every classification, the rule findings it identified (the skill’s triggers), the note type or section supporting each, a verbatim quotation from the source note, and the classification rule it applied. Where the engine judged that the record held nothing about alcohol it returned that statement in the field instead of quoted text, and those classifications are reported separately (Fig. 4f).

~~~
---
***name:*** *aud-adjudication*
***description:*** *>
Classify Alcohol Use Disorder (AUD) status from clinical chart notes using a six-point ordinal scale. Designed for LLM-based chart review that replicates expert hepatology fellow adjudication. Applicable to any EHR note corpus where AUD status needs structured extraction*.
***version:*** *"1.0"*
***license:*** *CC-BY-4.0
---*
~~~

~~~
# AUD Chart Adjudication — Guidance for the LLM Reviewer
~~~

~~~
This is a self-contained instruction set for an LLM (or a new human reviewer) classifying whether a patient has Alcohol Use Disorder (AUD) from their clinical chart. It is written to be followed literally and in order.
~~~

~~~
# 1. What You Are Being Asked to Do
~~~

~~~
You will be given the clinical chart documentation for one patient at a time. Your single job is to read the chart and assign exactly one label from a six-point scale describing how confident the evidence is that this patient has A lcohol Use Disorder.
~~~

~~~
You are not diagnosing the patient. You are reporting what the documentation supports. Think of yourself as a careful reader summarizing what the chart says about alcohol — not a doctor deciding what is true.
~~~

~~~
Three things must accompany every classification: (a) which evidence categories supported your decision, (b) any screening-tool scores you found (AUDIT/CA GE), and (c) a short supporting quote with where it came from (HPI, problem list, social history, etc.). Never give a label without the quote that justifies it.
~~~

~~~
# 2. Read This Before You Start
~~~

~~~
- Your task is to replicate what a careful chart reviewer would conclude. Follow the rules precisely, not your own clinical intuition.
- The review window matters. Focus on documentation most relevant to the clinical question. Evidence far outside the period of interest is context, not primary basis.
- One patient may have many documents. Read all of them for that patient before deciding. Evidence in any one note counts for the whole patient.
~~~

~~~
# 3. The Six-Point Scale
~~~

~~~
Assign exactly one:
~~~

~~~
- Definitely Yes: An explicit AUD diagnosis, documented alcohol dependence or abuse, alcoholic cirrhosis (the only alcohol-attributed disease that qualifies on its own), a positive AUDIT (>=8) or CAGE (>=2), OR documented alcohol withdrawal and its management (including CIWA monitoring). Any ONE of these is enough.
- Probably Yes: No standalone trigger — BUT the chart documents heavy/extensive prior drinking, problematic patterns, OR alcohol-attributed sequelae OTHER than cirrhosis (alcoholic hepatitis, alcoholic pancreatitis, ALD, Wernicke/Korsakoff). These rise to Definitely Yes only when combined with a corroborating trigger.
- Uncertain: Alcohol use is mentioned, but quantity, pattern, or severity can not be determined (e.g., "Alcohol: Yes" or "drinks daily" with no further detail).
- Probably No: Alcohol is mentioned only in the context of clearly social or occasional consumption.
- Definitely No: Documentation explicitly indicates the patient is a non-drinker, OR alcohol is explicitly ruled out as a contributor (e.g., "no history o f ETOH abuse," "cirrhosis not due to alcohol").
- Not Enough Data: No mention of alcohol use anywhere in the reviewed chart.
~~~

~~~
How these collapse for analysis: Definitely Yes + Probably Yes = "AUD Present." Definitely No + Probably No = "AUD Absent." Uncertain and Not Enough Data are handled separately.
~~~

~~~
## 3.1 The Definitely Yes Triggers — Memorize These
~~~

~~~
Any ONE of the following locks the classification at Definitely Yes. Nothing later in the chart can downgrade it.
~~~

~~~
1. Alcoholic cirrhosis named as such — Cirrhosis is the ONLY alcohol-attribut ed disease that is a standalone trigger.
2. Explicit AUD diagnosis or "alcohol dependence / alcohol abuse / alcoholism" in the narrative — Direct evidence.
3. Positive screening tool: AUDIT >=8 or CAGE >=2 — Validated instruments cro ssing their clinical threshold.
4. Documented alcohol withdrawal OR its management — including CIWA protocol, benzodiazepine taper for withdrawal, DTs, detox. CIWA placement counts EVEN IF the patient never manifests withdrawal signs.
~~~

~~~
# 4. The Principles You Must Follow
~~~

~~~
## 4.1 Narrative beats structured fields (most important rule)
~~~

~~~
When structured fields (e.g., Social History box "Alcohol: No") and provider narrative (HPI, Assessment/Plan, progress notes) disagree, the narrative wins. Structured fields are often auto-populated or carried forward unchanged. A structured "Alcohol: No" next to narrative "history of alcoholic cirrhosis" = Definitely Yes.
~~~

~~~
Exception: if a structured field is the ONLY evidence and there is no narrative about alcohol at all, accept the structured field at face value.
~~~

~~~
## 4.2 Current sobriety never lowers the classification
~~~

~~~
AUD is about whether the disorder exists or existed. "Sober since 2015," "in remission," "attends AA," and "quit drinking" are evidence that there was a disorder to recover from. If Definitely Yes criteria are met historically, the answer stays Definitely Yes.
~~~

~~~
## 4.3 One trigger is enough
~~~

~~~
Do not wait for corroboration. A single Definitely Yes trigger settles the classification.
~~~

~~~
## 4.4 Definitely No includes "ruled out as a contributor"
~~~

~~~
Use the broader definition: not only "patient is a non-drinker," but also any explicit statement ruling alcohol out — e.g., "no history of ETOH abuse," "cirrhosis not due to alcohol," "non-alcoholic steatohepatitis" when stated to exclude alcohol.
~~~

~~~
## 4.5 Classify only what is documented
~~~

~~~
Never infer a diagnosis the chart does not state. "Cirrhosis" with no stated cause is NOT automatically alcoholic.
~~~

~~~
# 5. How to Handle Marginal Cases
~~~

~~~
- Structured field conflicts with narrative → follow the narrative (Section 4.1).
- Single acute intoxication or "etoh fall" visit, no diagnosis/pattern/screen/withdrawal/sequelae → Probably Yes or Uncertain, NOT Definitely Yes. One event is not a disorder.
- Alcohol-attributed sequelae OTHER than cirrhosis (alcoholic hepatitis, pancreatitis, ALD, Wernicke/Korsakoff) WITHOUT a corroborating trigger → Probably Yes. They rise to Definitely Yes only when paired with another trigger.
- "Alcohol: Yes" or "drinks daily" with no quantity/pattern/severity → Uncert ain.
- Clearly social/occasional use → Probably No.
- Conflicting frequency entries in structured fields → let the clinical picture in the narrative decide.
- Patient sober/in remission but historical triggers present → Definitely Yes (Section 4.2).
- No mention of alcohol anywhere → Not Enough Data.
~~~

~~~
# 6. Common Mistakes and Corrections
~~~

~~~
- Trusting structured "Alcohol: No" over narrative → Narrative wins. Alcoholic cirrhosis in HPI = Definitely Yes regardless of structured field.
- Downgrading because patient is sober → Current abstinence is irrelevant. So ber 10 years + alcoholic cirrhosis = still Definitely Yes.
- Requiring several triggers → Any ONE trigger is sufficient.
- CIWA only counting if withdrawal occurred → CIWA initiation alone is the trigger.
- "Cirrhosis not due to alcohol" classified as Not Enough Data → This is Definitely No.
- Single intoxication = Definitely Yes → One acute event without diagnosis/pattern/withdrawal is Probably Yes or Uncertain.
- Inferring unstated diagnoses → Classify only what is documented. No alcohol mention = Not Enough Data.
~~~

~~~
# 7. Worked Examples
~~~

~~~
Example 1 — Structured-field trap: Social History "Alcohol: No" but HPI says "hx of alcohol related cirrhosis with portal HTN" → Definitely Yes. Narrative wins.
~~~

~~~
Example 2 — Clean denial: "Alcohol use: Denies." No alcohol-related anything → Definitely No.
~~~

~~~
Example 3 — CIWA without withdrawal: Patient on CIWA protocol, no formal AUD diagnosis → Definitely Yes. CIWA initiation is a standalone trigger.
~~~

~~~
Example 4 — Single intoxication: ER note for etoh fall, one event, no diagnos is/pattern → Probably Yes or Uncertain, NOT Definitely Yes.
~~~

~~~
Example 5 — Sober for years: "Alcoholic cirrhosis" + "sober since 2014, atten ds AA" → Definitely Yes. Sobriety never lowers.
~~~

~~~
Example 6 — Alcohol ruled out: "NASH cirrhosis. No history of ETOH abuse." → Definitely No.
~~~

~~~
Example 7 — Non-cirrhosis sequela alone: "Alcoholic pancreatitis" with no oth er triggers → Probably Yes (only cirrhosis is a standalone trigger).
~~~

~~~
Example 8 — Sequela + corroboration: "Alcoholic hepatitis" + "CIWA initiated"→ Definitely Yes. CIWA is a standalone trigger.
~~~

~~~
# 8. Final Checklist
~~~

~~~
Before submitting each classification:
- Did I read every document for this patient?
- Did I check whether any single Definitely Yes trigger is present?
- If structured and narrative disagree, did I follow the narrative?
- Did I avoid downgrading because the patient is currently sober?
- Is my label backed by an actual quote from the chart, with its source locat ion?
- Did I record any AUDIT/CAGE scores and the evidence categories I used?
- Am I classifying only what is documented — not what I assume?
~~~

**Supplementary Fig. 1.**
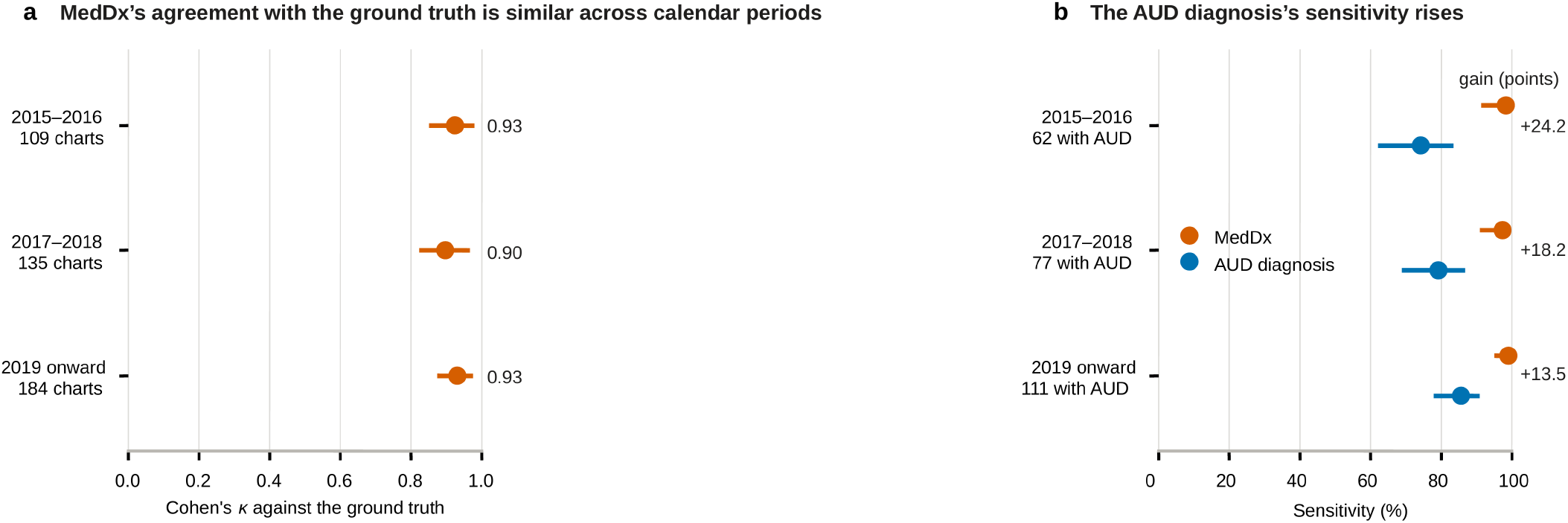
MedDx and the AUD diagnosis across calendar periods, on the reviewed charts. The bands are three calendar periods of the cirrhosis diagnosis (2015–16, 2017–18, 2019 onward) and contain 124, 150 and 211 charts; the 109, 135 and 184 printed in a are the charts both MedDx and the ground truth placed. **a**, Agreement between MedDx and the ground truth under the primary definition, with percentile bootstrap 95% CIs. **b**, Sensitivity of MedDx and of the AUD diagnosis on the ground-truth-positive charts in a band, with Wilson 95% CIs. MedDx is level across the three bands: κ 0.93, 0.90 and 0.93, and sensitivity 98%, 97% and 99%. The diagnosis moves: its sensitivity rises from 74% to 86% and the gain narrows from 24.2 points to 13.5. ICD-10-CM replaced ICD-9-CM in October 2015, inside the first band.

**Supplementary Fig. 2.**
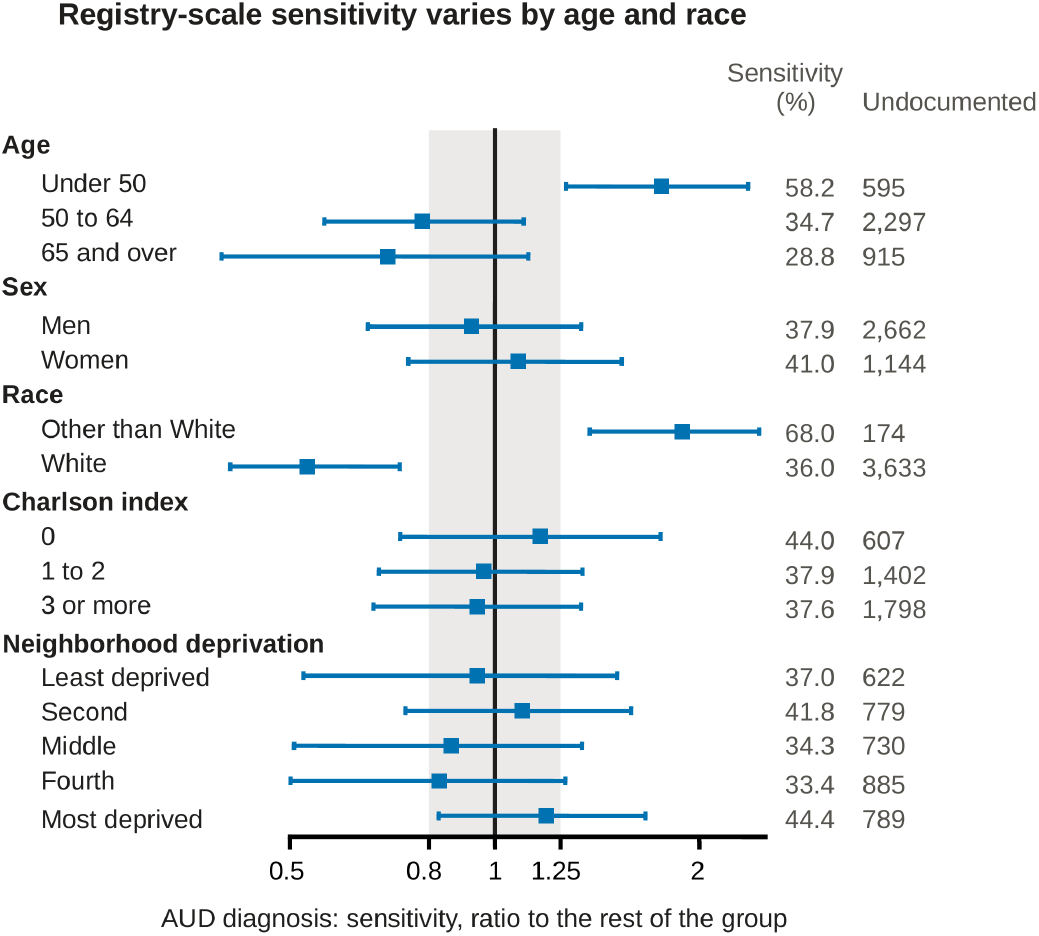
Registry-scale sensitivity of the AUD diagnosis within each patient group, as a ratio to the rest of the group. The registry estimate of the share of patients with AUD in each group who carry an AUD diagnosis at the cirrhosis diagnosis, weighted to the registry within hospital type and whole-record diagnosis status (Methods), divided by the same share in the remaining groups of the variable. One is no difference, on a logarithmic axis, and the gray band is 0.8 to 1.25. The columns give the group’s own sensitivity (%) and Undocumented, the estimated number of registry patients with AUD in the group who carry no AUD diagnosis. Intervals are 2,000-sample percentile bootstrap intervals over the AUD+ or AUD− charts, resampled within those cells (Supplementary Table 10). MedDx has no registry-scale counterpart here, because it read only the sampled charts; the chart-scale comparison is Fig. 3.

**Supplementary Fig. 3.**
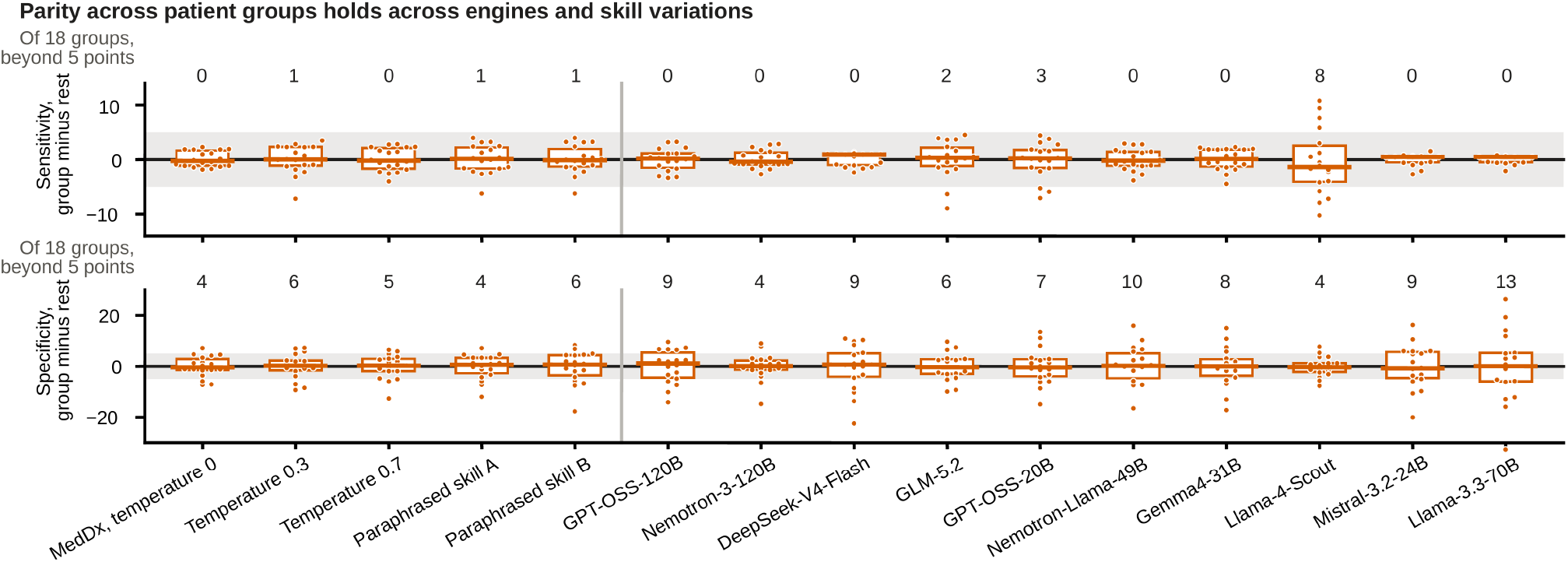
Sensitivity and specificity differences across patient groups for MedDx, its four skill variations and the other ten engines. Each point is one of the 18 groups of Fig. 3a and c: that group’s sensitivity (top) or specificity (bottom) minus the value in the rest of the group’s variable, in points. The box is the interquartile range, the line across it the median, the band is 5 points, and the row above each panel counts how many of that classifier’s 18 groups fall outside it. MedDx under the skill at temperature 0 and its four variations of Extended Data Fig. 5 are left of the rule; the other ten engines follow in the order of Fig. 4b. The AUD diagnosis is drawn beside MedDx in Fig. 3b. The widest specificity spreads belong to the classifiers that make the most errors (Supplementary Table 11).

**Supplementary Fig. 4.**
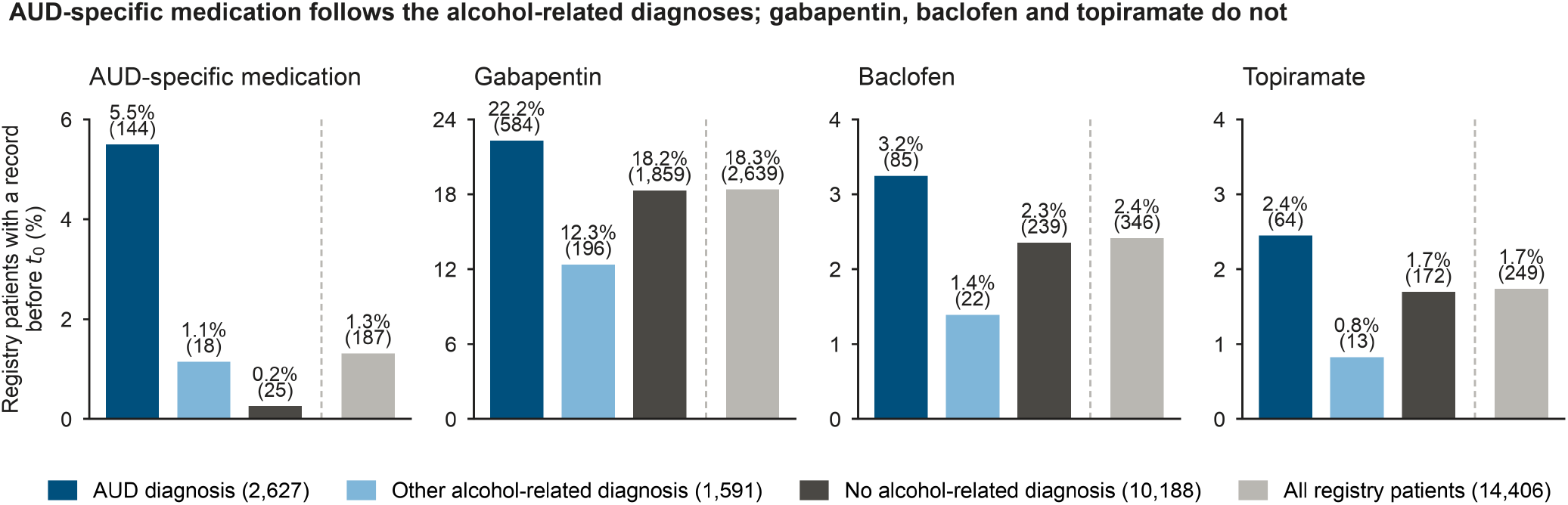
Medication recorded before the cirrhosis diagnosis, by stratum. Registry patients with a first record of the medication dated before the cirrhosis diagnosis, over the registry patients of each stratum; the bar right of the dashed line is the whole registry. AUD-specific medication is naltrexone, acamprosate or disulfiram, and its 187 patients are the prior users. Gabapentin, baclofen and topiramate are also prescribed for alcohol use disorder, but they are as common in the no alcohol-related diagnosis stratum as in the registry as a whole, so they are not counted as AUD treatment.

**Supplementary Table 1.** Six-class confusion matrix for MedDx against the ground truth. Rows are the ground truth, columns MedDx. *n* = 485. Of all classifications, 86.4% fall on the diagonal and a further 10.9% are one grade away on the ordinal scale. Not enough data has no position on that scale and is not counted in the second figure. The three-class collapse used throughout the main text is Fig. 1c.

| Ground truth | Def. Yes | Prob. Yes | Uncertain | Prob. No | Def. No | Not enough data |
|---|---|---|---|---|---|---|
| Def. Yes | 210 | 6 | – | – | – | – |
| Prob. Yes | 20 | 10 | 2 | 1 | 1 | – |
| Uncertain | 1 | 1 | 3 | 1 | 1 | – |
| Prob. No | 3 | 1 | 4 | 31 | 15 | 2 |
| Def. No | 1 | – | – | 4 | 112 | – |
| Not enough data | – | 1 | – | – | 1 | 53 |

**Supplementary Table 2.** Agreement with the ground truth within each grade of MedDx’s own scale. The denominator is the charts MedDx placed in each grade; agreement is with the ground truth collapsed to three classes. Intervals are Wilson intervals. The gradient runs in the expected direction on both sides: definite minus probable is +13.7 percentage points on the positive side and +3.1 on the negative side, with 19 probable-positive classifications.

| **Model classification** | **n** | **Agreement with the ground truth, % (95% CI)** |
|---|---|---|
| Def. Yes | 235 | 97.9 (95.1–99.1) |
| Prob. Yes | 19 | 84.2 (62.4–94.5) |
| Prob. No | 37 | 94.6 (82.3–98.5) |
| Def. No | 130 | 97.7 (93.4–99.2) |
| Uncertain | 9 | 33.3 (12.1–64.6) |
| Not enough data | 55 | 96.4 (87.7–99.0) |

**Supplementary Table 3.**
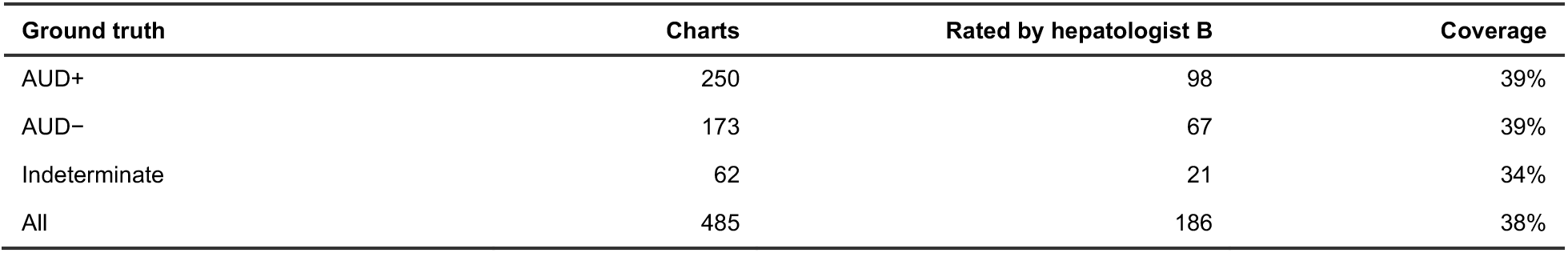
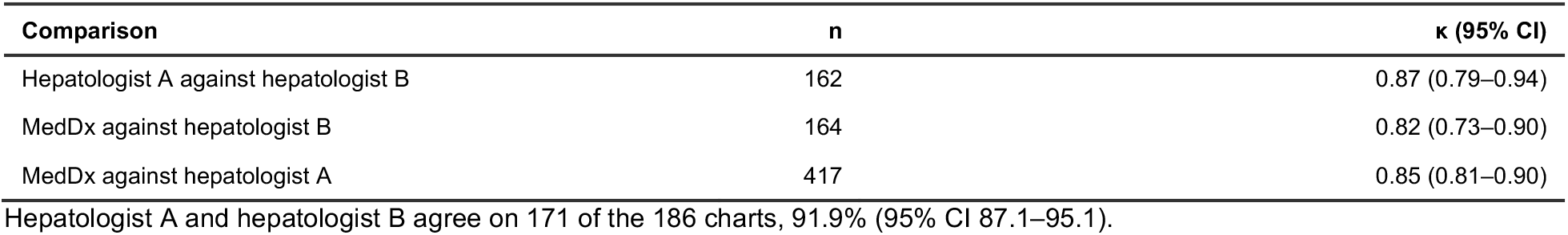
Hepatologist B’s independent review. Hepatologist B read the 186 charts of the study cohort, blind to hepatologist A’s classifications and to all MedDx output, and took no part in the adjudication. Coverage is over the 485 charts of the study cohort. Hepatologist B’s disagreement with hepatologist A is the second reason for a second reading, so every chart on which the two differ went to a second reading. The quantities below are computed over every chart hepatologist B read. Agreement is at the three-class level and κ uses the primary definition, three classes with not enough data excluded, both on raw first-pass classifications before any adjudication. The denominators of κ are therefore smaller than 186.

| Ground truth | Charts | Rated by hepatologist B | Coverage |
|---|---|---|---|
| AUD+ | 250 | 98 | 39% |
| AUD− | 173 | 67 | 39% |
| Indeterminate | 62 | 21 | 34% |
| All | 485 | 186 | 38% |
| **Comparison** | **n** |  | **κ (95% CI)** |
| Hepatologist A against hepatologist B | 162 |  | 0.87 (0.79–0.94) |
| MedDx against hepatologist B | 164 |  | 0.82 (0.73–0.90) |
| MedDx against hepatologist A | 417 |  | 0.85 (0.81–0.90) |

Hepatologist A and hepatologist B agree on 171 of the 186 charts, 91.9% (95% CI 87.1–95.1).

**Supplementary Table 4.** Sensitivity and specificity of MedDx on the reviewed charts, as sampled and weighted to the registry. Intervals are percentile bootstrap intervals.

| **Weighting** | **Quantity** | **Value, % (95% CI)** |
|---|---|---|
| As sampled | Sensitivity | 98 (97–100) |
|  | Specificity | 94 (90–97) |
| Weighted | Sensitivity | 98 (95–100) |
|  | Specificity | 94 (90–97) |

**Supplementary Table 5.** The nine cells of the registry estimate, hospital type by whole-record diagnosis status, with the registry patients and the reviewed charts in each. The Wilson 95% CI of each cell’s share with AUD is drawn in Extended Data Fig. 4. Hospital type comes from the sites of a patient’s inpatient and emergency visits, and diagnosis status is what the structured record holds at any time: an AUD diagnosis, an alcohol-related diagnosis other than AUD, or neither. The share with AUD is the ground-truth-positive share of the charts with an AUD+ or AUD− ground truth, and the patients with AUD are that share times the registry patients; the cells sum to 6,223 and the registry patients to 14,406. In the All rows the share is the patients with AUD over the registry patients.

| Diagnosis status at any time | Hospital type | Registry patients | Charts reviewed | Determinate | Ground-truth-positive | Share with AUD, % | Patients with AUD |
|---|---|---|---|---|---|---|---|
| AUD diagnosis | All | 4,210 | 233 | 219 | 202 | 92.6 | 3,900 |
|  | Academic health system | 2,328 | 130 | 125 | 116 | 92.8 | 2,160 |
|  | Safety-net hospital | 773 | 61 | 59 | 53 | 89.8 | 694 |
|  | Other hospitals | 1,109 | 42 | 35 | 33 | 94.3 | 1,046 |
| Other alcohol-related diagnosis | All | 1,957 | 49 | 43 | 32 | 77.0 | 1,507 |
|  | Academic health system | 876 | 21 | 19 | 14 | 73.7 | 645 |
|  | Safety-net hospital | 182 | 15 | 13 | 9 | 69.2 | 126 |
|  | Other hospitals | 899 | 13 | 11 | 9 | 81.8 | 736 |
| No alcohol-related diagnosis | All | 8,239 | 203 | 161 | 16 | 9.9 | 816 |
|  | Academic health system | 4,187 | 100 | 82 | 6 | 7.3 | 306 |
|  | Safety-net hospital | 523 | 23 | 23 | 3 | 13.0 | 68 |
|  | Other hospitals | 3,529 | 80 | 56 | 7 | 12.5 | 441 |
| All registry patients |  | 14,406 | 485 | 423 | 250 | 43.2 | 6,223 |

Of the 2,513 patients with AUD estimated in the no alcohol-related diagnosis stratum at *t*<sub>0</sub>, 816 (95% CI 473–1,290) are among the 8,239 never given an alcohol-related diagnosis, from 16 ground-truth-positive charts among the 161 reviewed charts with an AUD+ or AUD− ground truth; an estimated 832 are among the 918 who later received an AUD diagnosis and 866 among the 1,031 who later received another alcohol-related diagnosis.

**Supplementary Table 6.** Chart-scale difference between each patient group and the rest, in percentage points, for the AUD diagnosis and for MedDx. These are the values Fig. 3a and b draw. Charts is the number of ground-truth-positive and ground-truth-negative charts in the group; sensitivity is estimated on the first and specificity on the second. Sensitivity and specificity follow the convention of Fig. 1, under which an indeterminate output on a chart the ground truth placed counts against the classifier. Intervals are 4,000-sample percentile bootstrap intervals over charts, resampled within sampling stratum.

|  |  | AUD diagnosis |  | MedDx |  |
|---|---|---|---|---|---|
| Patient group | Charts | Sensitivity | Specificity | Sensitivity | Specificity |
| *Hospital system* |  |  |  |  |  |
| Academic | 120 / 71 | 9.7 (−0.0 to 19.4) | −0.1 (−9.1 to 8.6) | −1.7 (−5.1 to 1.1) | −1.2 (−8.7 to 6.2) |
| Safety net | 32 / 24 | −21.0 (−38.9 to −4.3) | 1.7 (−11.6 to 12.3) | 1.8 (0.4 to 3.8) | −7.1 (−23.0 to 5.4) |
| Other hospitals | 98 / 78 | −0.3 (−10.6 to 9.6) | −0.8 (−9.9 to 8.1) | 1.0 (−2.0 to 3.9) | 4.6 (−2.5 to 11.5) |
| Men | 165 / 69 | −5.9 (−15.9 to 4.1) | −0.5 (−10.0 to 8.5) | 1.1 (−1.9 to 4.9) | −1.5 (−9.5 to 5.8) |
| Women | 85 / 104 | 5.9 (−4.1 to 15.9) | 0.5 (−8.5 to 10.0) | −1.1 (−4.9 to 1.9) | 1.5 (−5.8 to 9.5) |
| Other than White | 33 / 27 | 4.7 (−9.5 to 17.3) | −5.9 (−21.7 to 7.1) | 1.8 (0.5 to 3.8) | −1.2 (−13.8 to 8.1) |
| White | 217 / 146 | −4.7 (−17.3 to 9.5) | 5.9 (−7.1 to 21.7) | −1.8 (−3.8 to −0.5) | 1.2 (−8.1 to 13.8) |
| Under 50 | 67 / 17 | 14.0 (4.7 to 23.1) | −15.2 (−37.4 to 3.8) | 0.1 (−3.7 to 3.2) | 7.1 (3.2 to 11.5) |
| 50 to 64 | 143 / 77 | −0.9 (−10.7 to 9.3) | −1.0 (−10.1 to 8.1) | −1.2 (−4.2 to 1.8) | −7.3 (−15.3 to 0.5) |
| 65 and over | 40 / 79 | −18.8 (−35.1 to −3.2) | 6.4 (−2.4 to 15.3) | 1.9 (0.5 to 4.0) | 4.7 (−2.5 to 11.8) |
| Urban | 184 / 114 | 4.8 (−6.2 to 16.8) | −9.7 (−17.3 to −1.7) | −0.1 (−3.1 to 3.8) | 3.4 (−4.6 to 12.2) |
| Suburban | 29 / 17 | 6.1 (−8.3 to 18.6) | 4.4 (−10.4 to 14.1) | 1.8 (0.4 to 3.7) | −6.0 (−24.1 to 7.5) |
| Rural | 37 / 41 | −12.4 (−28.6 to 2.9) | 9.8 (2.0 to 16.7) | −1.3 (−7.7 to 2.7) | −1.2 (−10.9 to 7.2) |
| Least deprived | 35 / 32 | −0.8 (−16.1 to 13.0) | 8.4 (−0.1 to 15.7) | −1.4 (−8.2 to 2.8) | −3.6 (−15.5 to 6.4) |
| Second | 51 / 32 | 2.1 (−10.5 to 13.6) | 4.5 (−6.0 to 13.3) | −0.4 (−5.3 to 3.0) | 0.2 (−10.1 to 8.5) |
| Middle | 43 / 35 | 0.9 (−13.4 to 13.6) | 1.7 (−10.5 to 11.3) | −0.9 (−6.5 to 2.8) | 0.9 (−8.6 to 8.6) |
| Fourth | 48 / 33 | −1.8 (−14.7 to 10.4) | 8.6 (−0.2 to 16.1) | −0.6 (−5.6 to 2.9) | 4.2 (−4.2 to 10.6) |
| Most deprived | 71 / 39 | −0.5 (−11.8 to 10.3) | −20.3 (−35.0 to −6.4) | 2.3 (0.5 to 4.7) | −1.6 (−11.4 to 6.9) |

**Supplementary Table 7.** The AUD diagnosis and MedDx by hospital system, on all 485 reviewed charts.

|  | **Academic health system** | **Other hospitals** | **Safety-net hospital** |
|---|---|---|---|
| Sensitivity of the AUD diagnosis, % | 85.8 | 80.6 | 62.5 |
| MedDx κ against the ground truth | 0.89 | 0.96 | 0.89 |
| Sensitivity gain, MedDx minus the diagnosis (percentage points) | +11.7 (+5.6 to +17.8) | +18.4 (+10.6 to +27.1) | +37.5 (+20.5 to +55.3) |

**Supplementary Table 8.** Supervised comparators trained at the academic health system and applied to the other hospital systems. The gradient-boosted model over the structured record, with the AUD diagnosis among its inputs, and the TF-IDF logistic regression over the notes were fitted on the 191 AUD+ or AUD− charts of the academic health system as specified in Supplementary Table 19, and applied unchanged to the safety-net hospital and the other hospitals. Academic-system rows are 5-fold out-of-fold estimates on the training charts. MedDx and the AUD diagnosis are shown on the same charts, and both are training-free. Wilson 95% CI. Fig. 3e shows this table.

| Classifier | Setting | Hospital system | Ground-truth-positive / negative charts | Sensitivity, % (95% CI) | Specificity, % (95% CI) |
|---|---|---|---|---|---|
| XGBoost, structured record with the AUD diagnosis | trained at the academic health system | Academic health system (training) | 120 / 71 | 94 (88–97) | 90 (81–95) |
| XGBoost, structured record with the AUD diagnosis | trained at the academic health system | Safety-net hospital | 32 / 24 | 81 (65–91) | 79 (60–91) |
| XGBoost, structured record with the AUD diagnosis | trained at the academic health system | Other hospitals | 98 / 78 | 89 (81–94) | 87 (78–93) |
| TF-IDF logistic regression, notes | trained at the academic health system | Academic health system (training) | 120 / 71 | 83 (76–89) | 72 (60–81) |
| TF-IDF logistic regression, notes | trained at the academic health system | Safety-net hospital | 32 / 24 | 94 (80–98) | 25 (12–45) |
| TF-IDF logistic regression, notes | trained at the academic health system | Other hospitals | 98 / 78 | 85 (76–90) | 51 (40–62) |
| AUD diagnosis (F10.x) | training-free | Academic health system (training) | 120 / 71 | 86 (79–91) | 90 (81–95) |
| AUD diagnosis (F10.x) | training-free | Safety-net hospital | 32 / 24 | 63 (45–77) | 92 (74–98) |
| AUD diagnosis (F10.x) | training-free | Other hospitals | 98 / 78 | 81 (72–87) | 90 (81–95) |
| MedDx | training-free | Academic health system (training) | 120 / 71 | 97 (93–99) | 93 (85–97) |
| MedDx | training-free | Safety-net hospital | 32 / 24 | 100 (89–100) | 88 (69–96) |
| MedDx | training-free | Other hospitals | 98 / 78 | 99 (94–100) | 96 (89–99) |

**Supplementary Table 9.**
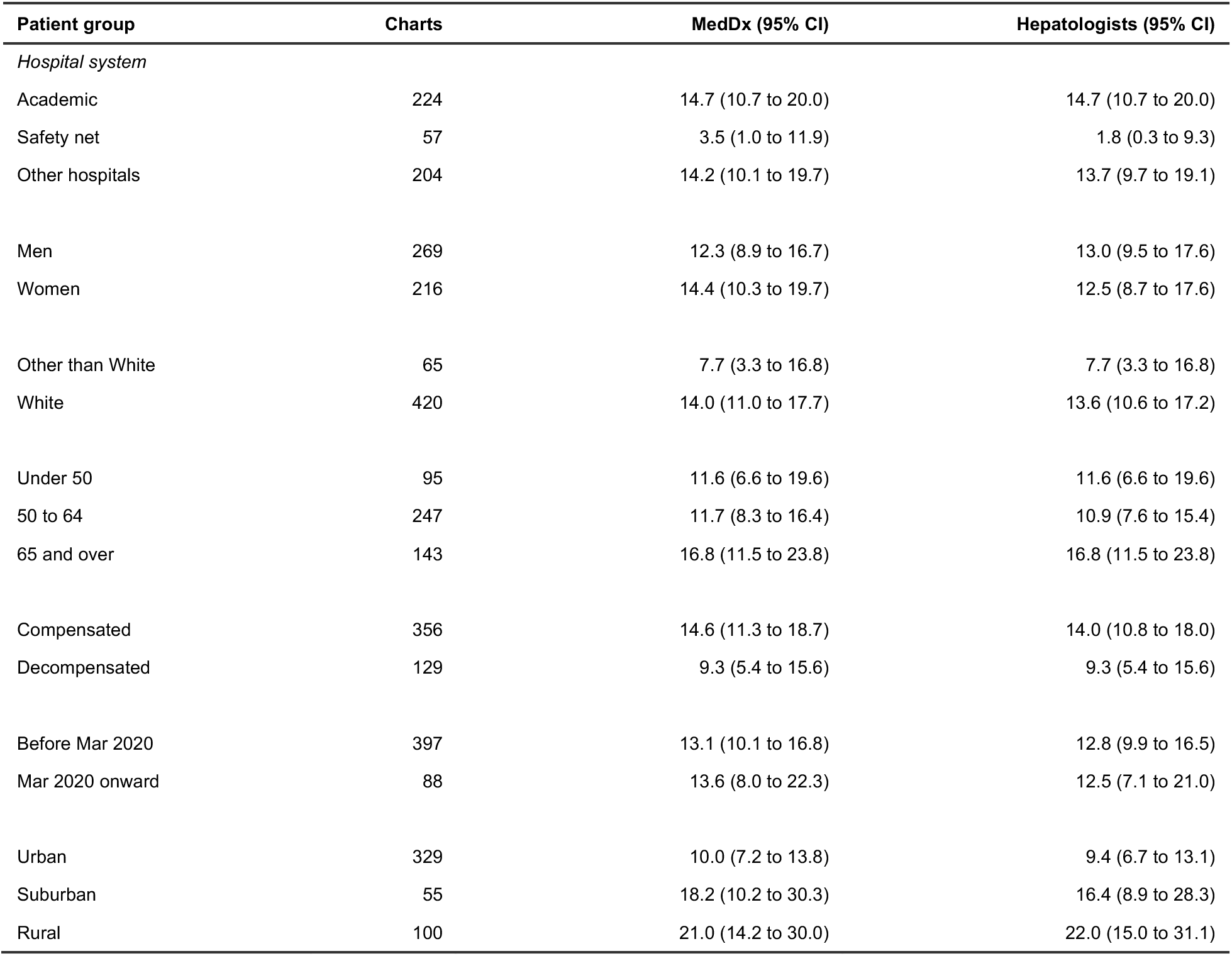
The rate at which an indeterminate classification is returned, by patient group, for MedDx and for the hepatologists on the same charts. The denominator is every chart in the group, whatever its ground-truth classification, so the quantity is not conditioned on the ground truth. Both indeterminate grades count on MedDx’s side, uncertain and not enough data, matching the single indeterminate class of the ground truth. Intervals are Wilson intervals. Charts is the number of charts the variable assigns to a level; one has no urban–rural classification.

**Supplementary Table 10.**
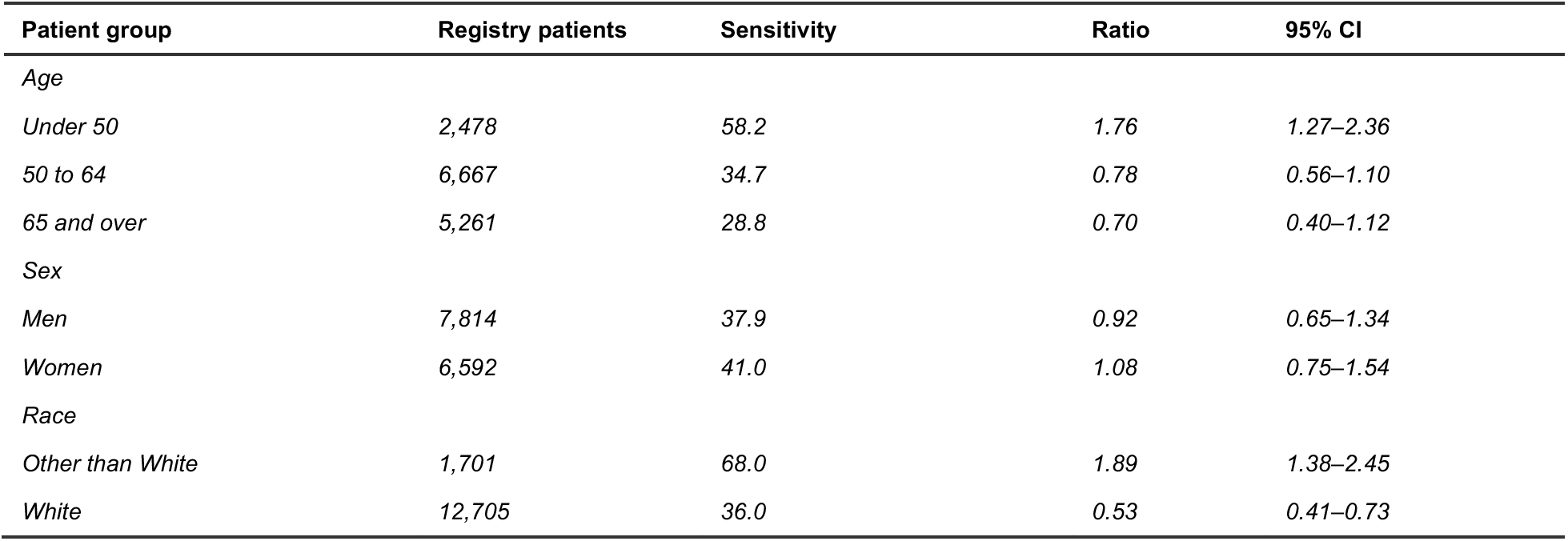
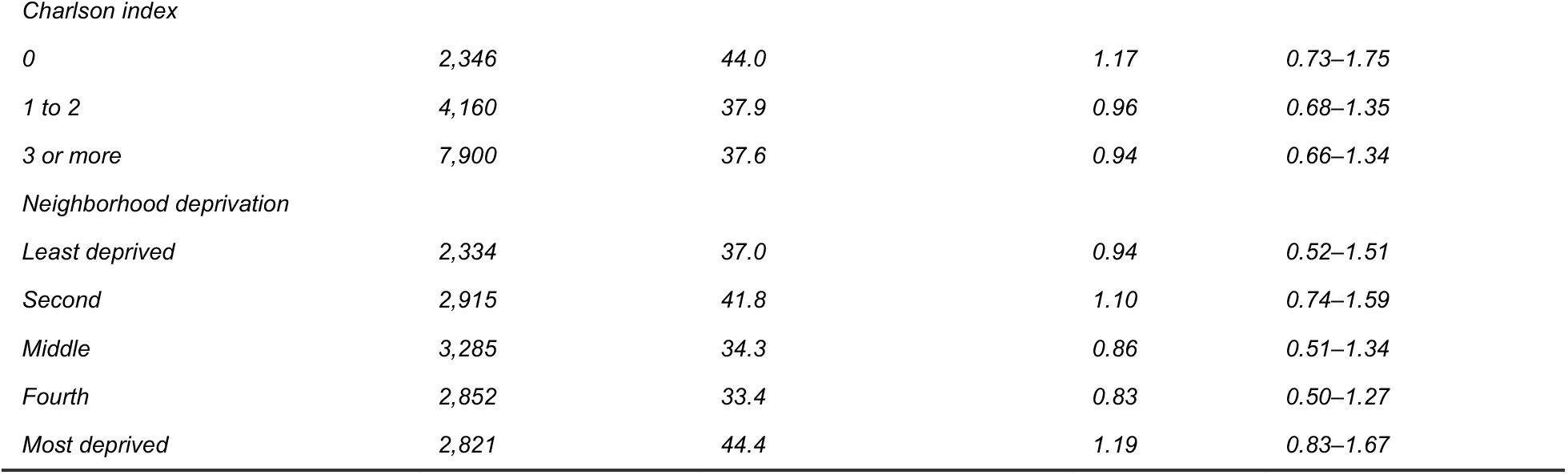
Registry-scale sensitivity of the AUD diagnosis by patient group, and its ratio to the rest. The estimand is the one Supplementary Fig. 2 shows: the share of registry patients estimated to have AUD in the group who carry an AUD diagnosis by the cirrhosis diagnosis, weighted to the registry within hospital type and whole-record diagnosis status, divided by the same share in the remaining levels of that variable. Intervals are 2,000-sample percentile bootstrap intervals over the AUD+ or AUD− charts, resampled within those cells. Registry patients is the number of registry patients in the group, not the number estimated to have AUD. The Charlson score excludes its two liver components because every registry patient has cirrhosis. Neighborhood deprivation is the Social Deprivation Index quintile of the patient’s ZIP code tabulation area, on population-weighted national bands.

**Supplementary Table 11.**
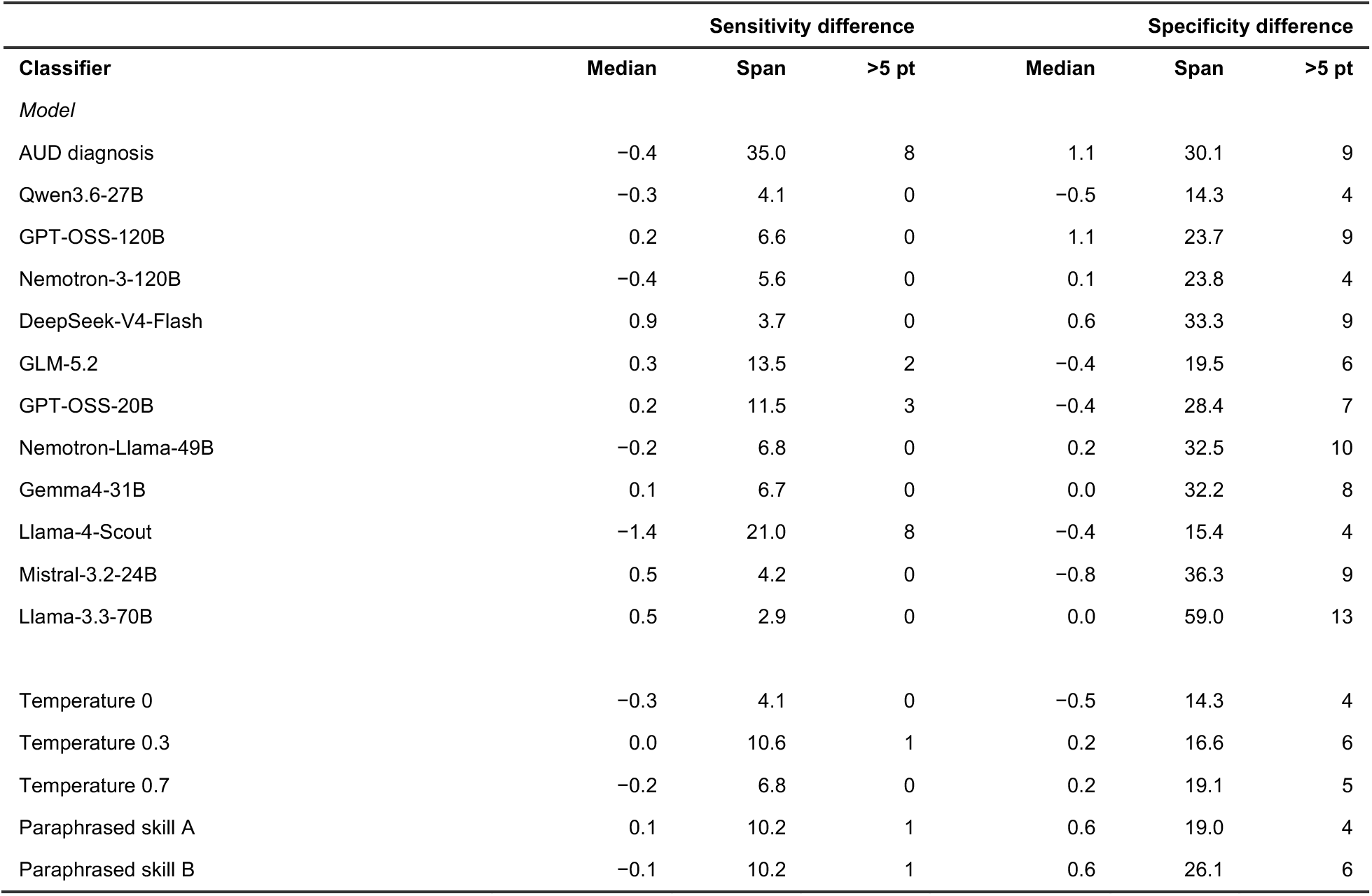
The spread of each classifier’s 18 group-minus-rest differences, on both metrics. Median and span are of the 18 differences and the last column of each pair counts those exceeding 5 points in absolute value. Every classifier’s median specificity difference lies between −0.8 and +1.1 points and the spans widen with the number of errors a classifier makes. The second block is the skill and its four variations, the five readings of Extended Data Fig. 5, by MedDx on the same 485 charts. Temperature 0 is MedDx as evaluated throughout.

| Classifier | Sensitivity difference |  |  | Specificity difference |  |  |
|---|---|---|---|---|---|---|
| Classifier | Median | Span | >5 pt | Median | Span | >5 pt |
| *Model* |  |  |  |  |  |  |
| AUD diagnosis | −0.4 | 35.0 | 8 | 1.1 | 30.1 | 9 |
| Qwen3.6-27B | −0.3 | 4.1 | 0 | −0.5 | 14.3 | 4 |
| GPT-OSS-120B | 0.2 | 6.6 | 0 | 1.1 | 23.7 | 9 |
| Nemotron-3-120B | −0.4 | 5.6 | 0 | 0.1 | 23.8 | 4 |
| DeepSeek-V4-Flash | 0.9 | 3.7 | 0 | 0.6 | 33.3 | 9 |
| GLM-5.2 | 0.3 | 13.5 | 2 | −0.4 | 19.5 | 6 |
| GPT-OSS-20B | 0.2 | 11.5 | 3 | −0.4 | 28.4 | 7 |
| Nemotron-Llama-49B | −0.2 | 6.8 | 0 | 0.2 | 32.5 | 10 |
| Gemma4-31B | 0.1 | 6.7 | 0 | 0.0 | 32.2 | 8 |
| Llama-4-Scout | −1.4 | 21.0 | 8 | −0.4 | 15.4 | 4 |
| Mistral-3.2-24B | 0.5 | 4.2 | 0 | −0.8 | 36.3 | 9 |
| Llama-3.3-70B | 0.5 | 2.9 | 0 | 0.0 | 59.0 | 13 |
| Temperature 0 | −0.3 | 4.1 | 0 | −0.5 | 14.3 | 4 |
| Temperature 0.3 | 0.0 | 10.6 | 1 | 0.2 | 16.6 | 6 |
| Temperature 0.7 | −0.2 | 6.8 | 0 | 0.2 | 19.1 | 5 |
| Paraphrased skill A | 0.1 | 10.2 | 1 | 0.6 | 19.0 | 4 |
| Paraphrased skill B | −0.1 | 10.2 | 1 | 0.6 | 26.1 | 6 |

**Supplementary Table 12.**
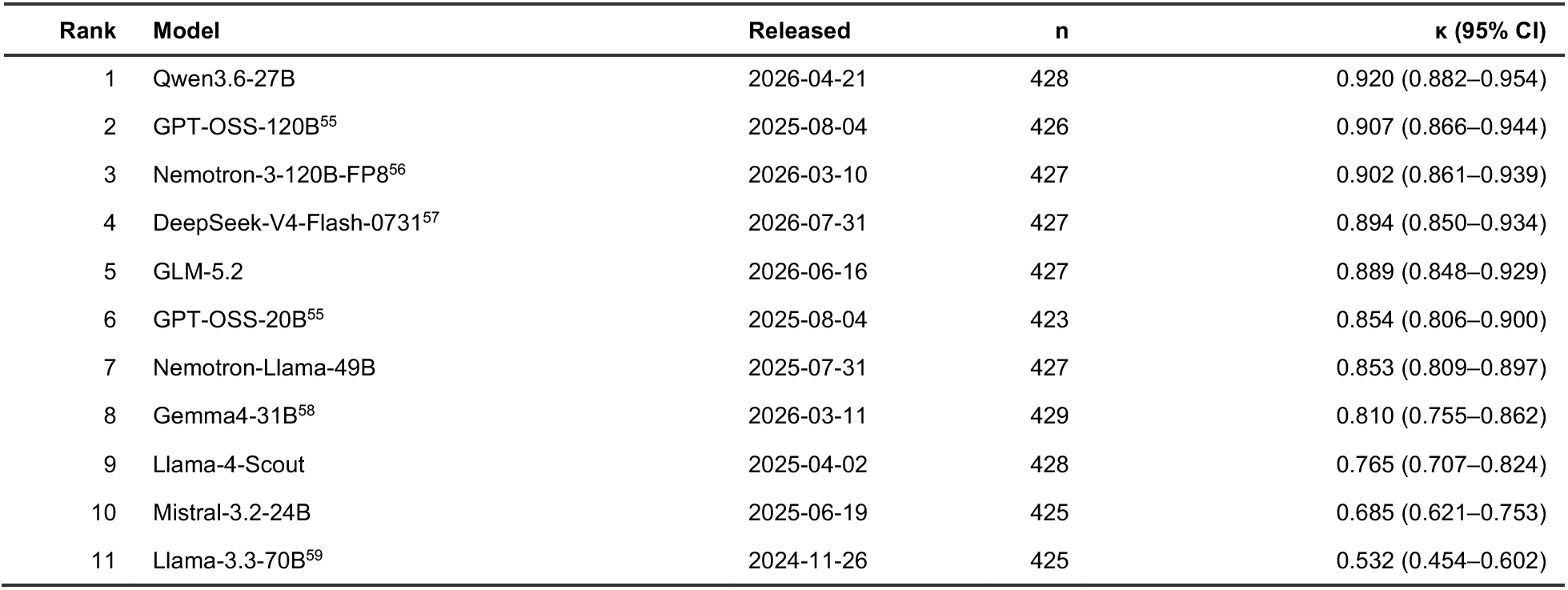
All 11 engines against the ground truth. Every engine processed the same 485 charts under the same skill, and no engine was adapted to the task. All are open-weight. The version string in the Model column identifies the weights, a citation marks each engine whose developer has published a report, and Released is the date the evaluated weights were first published (Supplementary Table 14). κ is the primary definition, three classes with not enough data excluded, so the denominator is the charts both classifiers placed and differs by engine (Methods). Intervals are 2,000-sample percentile bootstrap intervals. Fig. 4b shows the full ranking, and its stability is in Supplementary Table 13.

| Rank | Model | Released | n | κ (95% CI) |
|---|---|---|---|---|
| 1 | Qwen3.6-27B | 2026-04-21 | 428 | 0.920 (0.882–0.954) |
| 2 | GPT-OSS-120B<sup>55</sup> | 2025-08-04 | 426 | 0.907 (0.866–0.944) |
| 3 | Nemotron-3-120B-FP8<sup>56</sup> | 2026-03-10 | 427 | 0.902 (0.861–0.939) |
| 4 | DeepSeek-V4-Flash-0731<sup>57</sup> | 2026-07-31 | 427 | 0.894 (0.850–0.934) |
| 5 | GLM-5.2 | 2026-06-16 | 427 | 0.889 (0.848–0.929) |
| 6 | GPT-OSS-20B<sup>55</sup> | 2025-08-04 | 423 | 0.854 (0.806–0.900) |
| 7 | Nemotron-Llama-49B | 2025-07-31 | 427 | 0.853 (0.809–0.897) |
| 8 | Gemma4-31B<sup>58</sup> | 2026-03-11 | 429 | 0.810 (0.755–0.862) |
| 9 | Llama-4-Scout | 2025-04-02 | 428 | 0.765 (0.707–0.824) |
| 10 | Mistral-3.2-24B | 2025-06-19 | 425 | 0.685 (0.621–0.753) |
| 11 | Llama-3.3-70B<sup>59</sup> | 2024-11-26 | 425 | 0.532 (0.454–0.602) |

**Supplementary Table 13.** Rank of each engine by agreement with the ground truth over 2,000 resamples of patients: how often it comes first and the 2.5th to 97.5th percentile of its rank.

| **Engine** | **Quantity** | **Value** |
| **Engine** | **Quantity** | **Value** |
|---|---|---|
| Qwen3.6-27B | Comes first | 71.1%, rank 1 to 2 |
| GPT-OSS-120B | Comes first | 23.2%, rank 1 to 5 |
| DeepSeek-V4-Flash | Comes first | 4.8%, rank 1 to 6 |
| Nemotron-3-120B | Comes first | 0.9%, rank 2 to 5 |
| GLM-5.2 | Comes first | below 0.1%, rank 2 to 6 |
| All others | Comes first | below 0.1% |

**Supplementary Table 14.** The eleven LLM engines. Repository is the Hugging Face identifier of the exact weights evaluated. Released is the date those weights were first published there. Parameters gives the total count and, for mixture-of-experts models, the experts active per token. Weights is the numeric format in which the model was served, and FP8 (partial) marks a release that keeps some layers at higher precision. Rows are ordered by release date, newest first. Agreement for each engine is in Supplementary Table 12.

| Model | Hugging Face repository | Released | Parameters | Weights |
|---|---|---|---|---|
| DeepSeek-V4-Flash-0731 | huggingface.co/deepseek-ai/DeepSeek-V4-Flash-0731 | 2026-07-31 | 304B total; 6 experts per token | FP8 |
| GLM-5.2 | huggingface.co/zai-org/GLM-5.2 | 2026-06-16 | 753B total; 8 experts per token | FP8 |
| Qwen3.6-27B | huggingface.co/Qwen/Qwen3.6-27B | 2026-04-21 | 27.8B, dense | bfloat16 |
| Gemma4-31B | huggingface.co/google/gemma-4-31B-it | 2026-03-11 | 31.3B, dense | bfloat16 |
| Nemotron-3-120B-FP8 | huggingface.co/nvidia/NVIDIA-Nemotron-3-Super-120B-A12B-FP8 | 2026-03-10 | 124B total; 12B active | FP8 (partial) |
| GPT-OSS-120B | huggingface.co/openai/gpt-oss-120b | 2025-08-04 | 117B total; 4 of 128 experts | MXFP4 |
| GPT-OSS-20B | huggingface.co/openai/gpt-oss-20b | 2025-08-04 | 21.5B total; 4 of 32 experts | MXFP4 |
| Nemotron-Llama-49B | huggingface.co/nvidia/Llama-3_3-Nemotron-Super-49B-v1_5-FP8 | 2025-07-31 | 49.9B, dense | FP8 (partial) |
| Mistral-3.2-24B | huggingface.co/mistralai/Mistral-Small-3.2-24B-Instruct-2506 | 2025-06-19 | 24.0B, dense | bfloat16 |
| Llama-4-Scout | huggingface.co/meta-llama/Llama-4-Scout-17B-16E-Instruct | 2025-04-02 | 109B total; 17B active of 16 experts | bfloat16 |
| Llama-3.3-70B | huggingface.co/meta-llama/Llama-3.3-70B-Instruct | 2024-11-26 | 70.6B, dense | bfloat16 |

**Supplementary Table 15.** The alcohol-related diagnosis code set. Every ICD-10 and ICD-9 code counted as an alcohol-related diagnosis, both in the comparator of that name and in the definition of the registry strata. The first row is the AUD diagnosis itself, and the remaining rows are alcohol-related diagnoses other than AUD. A dash marks a condition with no code in that revision. Codes that are not specific to alcohol are excluded: ICD-9 577.0, which is acute pancreatitis of any cause, and the T51 and 980 codes for alcohols other than ethanol.

| **Condition** | **ICD-10** | **ICD-9** |
|---|---|---|
| Alcohol use disorder | F10 | 303, 305.0 |
| Alcohol-associated liver disease | K70 | 571.0–571.3 |
| Alcohol-induced pancreatitis | K85.2 | – |
| Alcoholic polyneuropathy | G62.1 | 357.5 |
| Alcoholic cardiomyopathy | I42.6 | 425.5 |
| Alcoholic gastritis | K29.2 | 535.3 |
| Alcohol-induced pseudo-Cushing syndrome | E24.4 | – |
| Alcohol-induced mental disorder | – | 291 |
| Toxic effect of ethanol or unspecified alcohol | T51.0, T51.9 | 980.0, 980.9 |
| Counseling for alcohol use | Z71.4 | – |
| History of alcohol misuse | – | V11.3 |

**Supplementary Table 16.** Sensitivity analyses of the registry estimate. Each row gives the number of registry patients estimated to have AUD, the prevalence, and the patients with AUD who carry no AUD diagnosis at any time, under one change to the primary estimate (Methods). The tipping-point rows give the indeterminate charts one quarter, one half and three quarters of the share with AUD among the AUD+ or AUD− charts of their cell; none of it is the lower bound and all of it is the primary estimate. The interval is given for the primary estimate only.

| Specification | Patients with AUD | Prevalence, % | With no AUD diagnosis at any time | Share of patients with AUD, % |
|---|---|---|---|---|
| Primary: indeterminate charts carry no weight | 6,223 (5,744–6,749) | 43.2 | 2,323 (1,863–2,819) | 37.3 |
| Indeterminate charts counted as AUD− | 5,564 | 38.6 | 1,944 | 34.9 |
| Indeterminate charts counted as AUD+ | 7,922 | 55.0 | 4,002 | 50.5 |
| Indeterminate charts at one quarter of their cell’s share with AUD | 5,729 | 39.8 | 2,039 | 35.6 |
| Indeterminate charts at one half of their cell’s share with AUD | 5,894 | 40.9 | 2,133 | 36.2 |
| Indeterminate charts at three quarters of their cell’s share with AUD | 6,058 | 42.1 | 2,228 | 36.8 |
| Six cells: hospital type by the AUD diagnosis | 6,195 | 43.0 | 2,295 | 37.0 |
| Hospital type of a reviewed chart from the source of its notes | 6,344 | 44.0 | 2,457 | 38.7 |

**Supplementary Table 17.** Agreement under seven definitions, for three comparisons. The first block is MedDx against the ground truth on the 485 charts of the study cohort. The second is hepatologist A against hepatologist B and the third MedDx against hepatologist B, both on the 186 charts hepatologist B read and both on raw first-pass classifications taken before any adjudication. The first row of the first block is the primary definition used throughout, unweighted κ on three classes with not enough data excluded. Every other row changes one element of it. n is the number of charts the row’s definition scores, and differs across rows because some definitions exclude a classifier’s abstentions. Agreement is exact agreement on the classes the row defines. † A range over the six positions not enough data can occupy on the ordinal scale, not a bootstrap interval.

| Definition | n | κ (95% CI) | Agreement |
|---|---|---|---|
| Three classes, not enough data excluded *(primary)* | 428 | 0.92 (0.88–0.95) | 96.0% |
| Three classes, not enough data kept as indeterminate | 485 | 0.93 (0.90–0.96) | 95.7% |
| Three classes, linear weights | 485 | 0.94 (0.91–0.97) | 95.7% |
| Three classes, Gwet’s AC1 | 485 | 0.94 (0.91–0.96) | 95.7% |
| Two classes, both classifiers committed | 415 | 0.96 (0.94–0.99) | 98.3% |
| Five classes, linear weights | 428 | 0.90 (0.88–0.93) | 85.5% |
| Six classes, linear weights | 485 | 0.89–0.92† | 86.4% |
| Three classes, not enough data excluded *(primary)* | 162 | 0.87 (0.79–0.94) | 93.2% |
| Three classes, not enough data kept as indeterminate | 186 | 0.87 (0.80–0.93) | 91.9% |
| Three classes, linear weights | 186 | 0.89 (0.82–0.94) | 91.9% |
| Three classes, Gwet’s AC1 | 186 | 0.88 (0.82–0.94) | 91.9% |
| Two classes, both classifiers committed | 154 | 0.93 (0.87–0.99) | 96.8% |
| Five classes, linear weights | 162 | 0.89 (0.83–0.93) | 85.8% |
| Six classes, linear weights | 186 | 0.86–0.88† | 84.9% |
| Three classes, not enough data excluded *(primary)* | 164 | 0.82 (0.73–0.90) | 90.9% |
| Three classes, not enough data kept as indeterminate | 186 | 0.82 (0.74–0.89) | 89.2% |
| Three classes, linear weights | 186 | 0.84 (0.76–0.90) | 89.2% |
| Three classes, Gwet’s AC1 | 186 | 0.85 (0.78–0.91) | 89.2% |
| Two classes, both classifiers committed | 156 | 0.88 (0.80–0.95) | 94.2% |
| Five classes, linear weights | 164 | 0.85 (0.78–0.92) | 87.8% |
| Six classes, linear weights | 186 | 0.83–0.86† | 86.6% |

The comparison in the last two blocks is the one the first Results subsection rests on, and the intervals of the two overlap in every row that has one.

**Supplementary Table 18.** Every comparator against the ground truth. All eight classifiers were scored on the same 423 charts with an AUD+ or AUD− ground truth, and an indeterminate output stays in the denominator of sensitivity and specificity. Sensitivity and specificity have Wilson 95% confidence intervals. Predictive values are over the charts a classifier placed as AUD+ or AUD−, at the prevalence of the study cohort, and are not weighted to the registry. κ is on the two directional classes, over the 423 charts for the comparators, which always return one, and over the 415 on which it committed for MedDx, so it differs from the three-class κ of the main text. Each comparator’s feature set, tuning grid and resampling scheme is in Supplementary Table 19.

| Comparator | Sensitivity, % (95% CI) | Specificity, % (95% CI) | PPV, % | NPV, % | κ |
|---|---|---|---|---|---|
| MedDx | 98 (96–99) | 94 (89–96) | 98 | 99 | 0.96 |
| AUD diagnosis (F10.x) alone | 81 (76–85) | 90 (85–94) | 92 | 76 | 0.69 |
| Any alcohol-related diagnosis | 94 (90–96) | 84 (78–89) | 89 | 90 | 0.78 |
| AUD or alcohol-associated liver disease diagnosis | 94 (90–96) | 84 (78–89) | 90 | 90 | 0.79 |
| Gradient boosting over structured data, with the AUD diagnosis | 90 (86–94) | 86 (80–90) | 90 | 86 | 0.77 |
| Gradient boosting over structured data, without the AUD diagnosis | 90 (85–93) | 85 (79–90) | 90 | 85 | 0.75 |
| TF-IDF logistic regression, out of fold | 81 (75–85) | 73 (66–79) | 81 | 72 | 0.54 |
| Fine-tuned Bio_ClinicalBERT, out of fold | 82 (76–86) | 83 (77–88) | 88 | 76 | 0.64 |

**Supplementary Table 19.** How each comparator was built. Reads names the part of the record the comparator sees. Features or match rule gives its inputs, Fitting and tuning the estimator, its hyperparameter grid and its seed, and Decision rule how a score becomes a classification. The three code-based rows require no fitting. The two supervised models over the notes were fitted and evaluated out of fold on the 423 charts with an AUD+ or AUD− ground truth, with hyperparameters selected in an inner loop. Full metrics for every row are in Supplementary Table 18.

| Comparator | Reads | Features or match rule | Fitting and tuning | Decision rule |
|---|---|---|---|---|
| MedDx | Notes | Each patient’s notes, under the skill | No fitting; greedy decoding at temperature 0 | Six grades, collapsed to three |
| AUD diagnosis (F10.x) alone | Structured | The AUD diagnosis | None | Diagnosis present |
| Any alcohol-related diagnosis | Structured | Every alcohol-related diagnosis code, not F10.x alone: alcohol-associated liver disease, alcoholic pancreatitis, gastritis, neuropathy and the rest of Supplementary Table 15 | None | Any code in the set present |
| AUD or alcohol-associated liver disease diagnosis | Structured | The AUD diagnosis or alcohol-associated liver disease | None | Either present |
| Gradient boosting over the structured record | Structured | Age, sex, race, ethnicity, the 19 Charlson conditions, the comorbidity index, encounter count, note count, and two alcohol-code flags. Hospital system is not a feature. Fitted with and without the AUD diagnosis | XGBoost, histogram tree method, logistic loss. Grid over maximum depth {2, 3}, trees {150, 300} and learning rate {0.05, 0.1}, selected on the inner-loop area under the receiver operating characteristic curve. Seed 20260809 | Score at or above the training fold’s prevalence quantile |
| TF-IDF logistic regression over the notes | Notes | Word and bigram counts on each patient’s concatenated notes, sublinear term frequency with inverse document frequency, minimum document frequency 3, at most 200,000 features, lower-cased | L2-regularized logistic regression, inverse regularization strength over {0.01, 0.1, 1, 10} selected in the inner loop, at most 2,000 iterations. Seed 20260810 | Score at or above the training fold’s prevalence quantile |
| Fine-tuned Bio_ClinicalBERT over the notes | Notes | emilyalsentzer/Bio_ClinicalBERT<sup>30</sup>, a BERT encoder pretrained on clinical notes. Each patient’s concatenated notes are divided into non-overlapping 512-token windows, each inheriting the patient’s ground-truth classification. Training uses at most 32 windows per patient, at even spacing; inference scores every window | Fine-tuned for sequence classification. AdamW, weight decay 0.01, batch 16, three epochs, linear schedule with 10% warmup, gradient clipping at 1.0, bfloat16. Learning rate over {2 × 10<sup>−5</sup>, 5 × 10<sup>−5</sup>} selected in the inner loop. Seed 20260810 | Mean window score at or above the training fold’s prevalence quantile |

